# Interactions between influenza and respiratory syncytial viruses: two-pathogen modelling to analyse multiplex data

**DOI:** 10.64898/2026.09.03.26362151

**Authors:** George Shirreff, Sandra S. Chaves, Laurent Coudeville, Beatriz Mengual-Chuliá, Ainara Mira-Iglesias, Joan Puig-Barberà, Alejandro Orrico-Sanchez, Javier Díez-Domingo, Valencia Hospital Surveillance Network for the Study of Influenza and Other Respiratory Viruses (VAHNSI), F. Xavier Lopez-Labrador, Lulla Opatowski

**Affiliations:** Epidemiology and Modelling of Antibiotic Evasion (EMAE), Institut Pasteur, Université Paris Cité, Paris, France; Anti-infective evasion and pharmacoepidemiology team, Université Paris-Saclay, UVSQ, Inserm, CESP, Montigny-Le-Bretonneux, France; Centre of Excellence in Respiratory Pathogens (CERP), Hospices Civils de Lyon (HCL) and Centre International de Recherche en Infectiologie (CIRI), Équipe Santé Publique, Épidémiologie et Écologie Évolutive des Maladies Infectieuses (PHE3ID), Inserm U1111, CNRS UMR5308, ENS de Lyon, Université Claude Bernard Lyon 1 (UCBL Lyon 1), Lyon, France; New Products & Innovation, Sanofi Vaccines, Lyon, France; CIBER de Epidemiología y Salud Pública, Instituto de Salud Carlos III, Madrid, Spain; Virology Laboratory, Genomics and Health Area, Fundación para el Fomento de la Investigación Sanitaria y Biomédica de la Comunitat Valenciana, (FISABIO-Public Health), Valencia, Spain; Vaccines Research Unit, Fundación para el Fomento de la Investigación Sanitaria y Biomédica de la Comunitat Valenciana, (FISABIO-Public Health), Valencia, Spain; Universidad Católica de Valencia San Vicente Mártir, Valencia, Spain; Department of Microbiology and Ecology, Medical School, University of Valencia, Valencia, Spain

**Author notes:** Corresponding author: George Shirreff, Université Claude Bernard Lyon 1, Lyon, France. The Network members are listed in the Acknowledgements. F.X.L.-L. and L.O. contributed equally to the manuscript.

**Keywords:** Within-host interaction, viral interaction, influenza, respiratory syncytial virus, two-pathogen model, deterministic model, Bayesian inference, multiplex analysis, hospital surveillance, influenza-like illness

## Abstract

**Background:** Influenza and respiratory syncytial virus (RSV) both cause annual peaks in hospitalisation and with considerable overlap. Epidemiological studies suggest an inhibitory relationship, which is supported by laboratory studies. Several biological mechanisms including resource competition and immune activation have been identified, but limited evidence exists to distinguish their effects at the population level.

**Methods:** We developed an influenza-RSV compartmental model to estimate and compare potential interactions using data on virus-associated hospitalisations in Valencia, Spain from 2010 through 2020. We considered five interaction mechanisms: altered susceptibility, transmissibility and severity, a refractory period post-infection, and a residual positivity period. Using Bayesian inference, we fit the model to weekly hospitalisations with influenza, RSV or both, and to national surveillance data for influenza-like illness.

**Results:** We found evidence for inhibition between influenza and RSV, with several interaction mechanisms being supported. In the best-fitting model, active RSV infection reduced the probability of hospitalisation with influenza to 20% of the baseline (with the data supporting values from 0%-60%). Compared to a model without interaction, this mechanism reduced the number of hospital cases with influenza-RSV co-detection by 63%.

**Conclusions:** Our results support the existence of inhibitory mechanisms, with small but substantial effects on disease epidemiology. These results highlight the importance of understanding these interactions under the changing epidemiology of respiratory viruses and vaccinations.

**Summary:** We identified potential mechanisms of within-host interaction between influenza and respiratory syncytial virus (RSV) using two-pathogen models and a 10-year active surveillance cohort. In the best supported model, active RSV infection reduced the hospitalisation probability of influenza infection.

## Background

Influenza and respiratory syncytial virus (RSV) have overlapping transmission seasons in the temperate northern hemisphere^1^, presenting opportunities for co-infection. However, it is hypothesised that these viruses interact competitively at a population-level ^2^.

The body of literature documenting interactions between respiratory viruses, particularly with influenza, is growing ^3–5^. Influenza and RSV interactions are among the best studied it remains a complex picture. At the cellular level, they can inhibit one another through direct competition for resources within the same cell^6^ or the protective effect of extracellular interferons ^7^. Mouse studies have demonstrated that influenza can induce non-specific protective immune activation ^8^, inhibit RSV replication ^9^ and reduce morbidity during subsequent RSV infection ^10^, while RSV can reduce morbidity during subsequent infection with influenza ^11,12^. Human clinical studies have suggested that interaction between influenza and RSV may reduce co-circulation by inhibiting viral load titres ^13^, or have no effect on infection severity ^14^.

However, these results show inconsistencies, asymmetric effects and sometimes deleterious interactions. A study in which mice were initially infected with influenza A and then RSV found lower RSV titres but more severe infection compared with influenza-naïve mice ^15^. An observational study of influenza and RSV in hospitalised patients found that co-infection was associated with greater severity ^16^, with compounding of pulmonary congestion offering a potential mechanism^15^.

Several studies have reported on potential interactions by counting simultaneous detections (codetections) and comparing this with numbers expected under independent co-circulation ^4,17–21^. They have generally found that influenza and RSV co-detections are observed less often than expected, suggesting a negative interaction. However, analyses with Bayesian autoregressive models which control for patient characteristics and time correlations have found only weak correlations ^22–24^.

Mechanistic modelling studies which explicitly account for infection, co-infection and hospitalisation can formalise possible interaction mechanisms and challenge them with data. In this study we developed a two-pathogen model to identify such mechanisms between influenza and RSV in either direction.

## Methods

### Study population

The Valencia Hospital Surveillance Network for the Study of Influenza and Other Respiratory Viruses (VAHNSI) is an active surveillance network prospectively analysing respiratory hospitalisations in tertiary-care public hospitals in the Valencia Region of Spain ^25^. The current analysis was conducted over 2010-2020. Potential participants were those of all ages admitted to participating hospitals with acute respiratory illness. Informed consent was taken before enrolment from patients, or their legal guardians where appropriate ^25^. Samples were analysed using a multiplex polymerase chain reaction (PCR) panel that included RSV and influenza viruses ^26,27^. A co-detection sample is defined as one positive for both viruses.

### National surveillance data

The rates of medical care for influenza-like illness (ILI) was used as a proxy for all respiratory infections occurring within the community. We used weekly national incidence rates of ILI for Spain, from the WHO Global Influenza Programme database FluID from FluMart ^28,29^ via the European Centres for Disease Control ^30^. This was defined as the rate of patients with an acute respiratory infection, beginning within the previous 10 days, with cough and a fever of ≥38°C.

### Model

The population dynamics of infection and co-infection with two viruses were modelled using a modified deterministic compartmental SEIR model with age structure. The possible infection states included: Susceptible (S); latently infected (E); Infectious (I); and recovered. The recovered state, in which an individual is no longer infectious nor susceptible to the recovered virus, was separated into a potential positive testing period (Rv) during which the individual would continue to test positive for the recovered virus; a potential refractory period (Rf) in which the individual tests negative but is temporarily protected from infection with the other virus; and full recovery (R) in which the individual is no longer protected from the other virus. In the two-virus model, an individual can be in any combination of states for each virus (Figure 1a).

**Figure 1.**
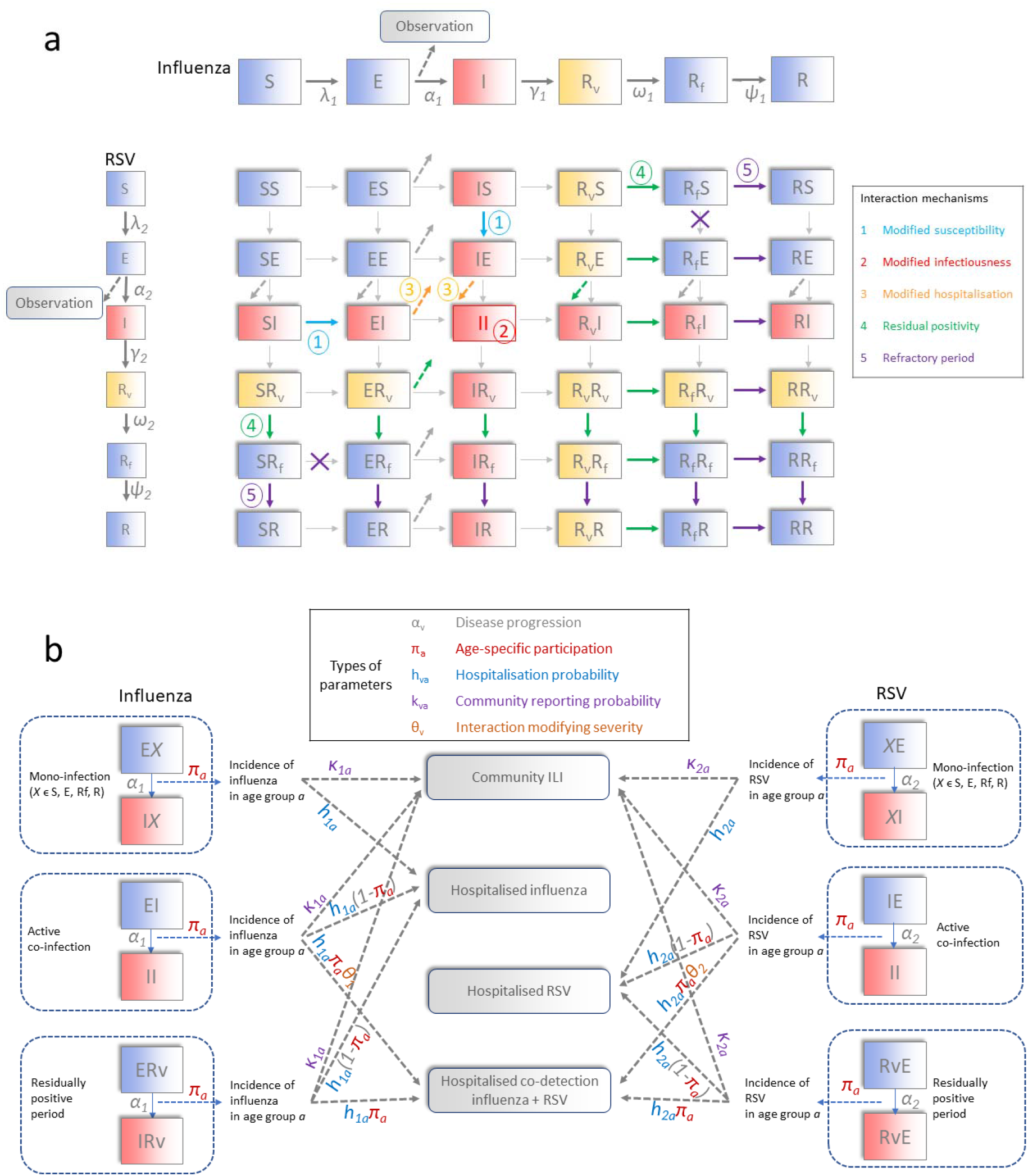
a) Schematic of model states across two viruses. The SEIRvRfR models on the top and left shown the progression of states of infection for influenza and respiratory syncytial virus (RSV) respectively. In the two-state model individuals can be in any state for either virus. The boxes show the possible modelled states, the vertical and horizontal arrows show paths of progression, and the coloured arrows show processes which are hypothesised as possible routes of interaction, which are shown and numbered in the table. b) Schematic of the observation process. The boxes on the far left and right represent instances of progression to active infection (E to I) for influenza and RSV, respectively. The rates of progression are given by, and the proportion becoming an incident infection given by for each age group a. The boxes in the centre represent the possible reporting outcomes, with the dotted arrows representing the path to each observation, with the parameter values indicating its probability. [Model states: S=susceptible; E=latently infected; I=infectious; Rv=recovered but testing positive; Rf=recovered refractory; R=recovered – Model parameters for each virus v: =force of infection; =progression rate from latency; =recovery rate from active infection; =progression rate from residual positive period; =progression rate from refractory period; =progression rate from refractory period; =hospitalisation probability for age group a; =community ILI consultation probability for age group a; =probability of participation in the epidemic for age group a; =modification of probability of hospitalisation with virus v by active infection with the other virus] Alt text: Schematic diagrams showing the construction of the two-pathogen model, with part a representing disease states with influenza and RSV, and part b representing the observation process.

We modelled each season as a separate event, assumed no waning immunity ^31^, and that the epidemic occurred in all age groups simultaneously, but allowing for age-specific probabilities of participating in the epidemic by becoming infected. At the beginning of each season, the entire population was assumed susceptible to both viruses, before an importation of each virus. The force of infection is calculated by multiplying the transmission rate for each virus and season by the proportion of infectious individuals (with interaction-modified transmission if appropriate).

Progression rates between stages of infection are virus-specific (Figure 1a). A full list of model parameters and their default values is in Supplementary Table 1.

### Virus interaction mechanisms

The following mechanisms were evaluated, each operating at different stages of infection (Figure 1a): (1) modified susceptibility, whereby those in active infection (I) with virus *y have their* susceptibility to virus *v multiplied* by a factor *δ*_*V*_, *affectin*g their infection from S to E; (2) modified infectiousness, whereby for those in the compartment with two active infections (II), the contribution to the force of infection for each virus is multiplied by a factor *σ* _*V*_; (3) modified severity, whereby those in active infection (I) with virus *y have their* probability of hospitalisation upon progression to active infection (from E to I) of virus v multiplied by a factor θV; (4) allowing for a period of residual positivity by extending the duration of the Rv stage *(1⁄ω*_*V*_*); or (5)* allowing for a period of refractory protection by extending the duration of the Rf stage *(1⁄ψ*_*V*_*). Importa*ntly, the Rv and Rf stages were only ever considered separately, meaning one or both always had negligeable length in the same analysis.

Interaction mechanisms were assumed to operate uniformly across all age groups.

### Observation process

Average contact rates by age group were estimated from the POLYMOD survey data ^32^ and standardised relative to the highest contact age group (5-17 years) to give the age-specific relative probability of participating in the epidemic, *π*_*a*_.

*Infectio*ns could be observed upon progression to infectiousness, and recorded as community ILI, hospitalisation with a single virus, or hospitalisation with co-detection, according to the participation probability *π*_*a*_, *the hosp*italisation probability (*h*_*Va*_*), and th*e probability of ILI consultation (*k*_*Va*_) (Figure 1b). The likelihood of observing the number of hospitalised cases in each week, age group and virus or co-detection given the model was calculated, as well as the likelihood of observing the number of community ILI by week and age group.

Model equations and further details of the methods are provided in the Supplementary Information, describing the evolution of the infectious status of the population (Model state equations), the process of observation for the hospitalisation and ILI consultation data (Model observation equations), and the likelihood calculation (Likelihood equations). The evolution of the internal state of the model was implemented using the *odin*^*33*^ *library in R*^*34*^.

### *Model* Inference

The two-virus time series were analysed jointly for each season independently, because the timing varied in each seasonal epidemic. The transmission rates *β*_*VS*_ *and week* of introduction *η*_*VS*_ *were est*imated for each virus *v and season s. The hospi*talisation rates *h*_***va***_ were estimated over the entire study period for each virus *v and age gro*up *a. Similarly* the community ILI consultation probabilities *k*_*Vb*_ *were est*imated over the whole period for each virus *v and age gro*up *b*.

*Parameter*s were estimated using adaptive Markov Chain Monte Carlo (MCMC) methods from the from the *FME R package*^35^. First, the baseline model was estimated, assuming no interaction, meaning that all interaction parameters were fixed to baseline values, while the non-interaction parameters (transmission rate, season start time, hospitalisation probability and community ILI consultation probability) were estimated. The best Akaike Information Criterion (AIC) value was calculated for the baseline model (with K=66 parameters) based on the highest likelihood.

Secondly, we fit models in which non-interaction parameters were fixed and a single interaction parameter or pair thereof was allowed to vary, to assess where this improved the fit to the data. Model ability to improve on the baseline model was based on AIC comparison, and we report interaction parameter values which resulted in smaller AIC than the baseline model. Parameter value estimates were not provided for models which did not improve the fit. The best-fit model was the one with the highest posterior likelihood value.

Further details are given in the Supplementary Information (Model inference process).

## Results

Weekly incidence of influenza virus and RSV from hospital surveillance in Valencia shows the extent of yearly co-circulation, consistent with winter peaks in national ILI incidence (Figure 2). Hospitalisation data is shown for finer age categories in Supplementary Figure 1.

**Figure 2.**
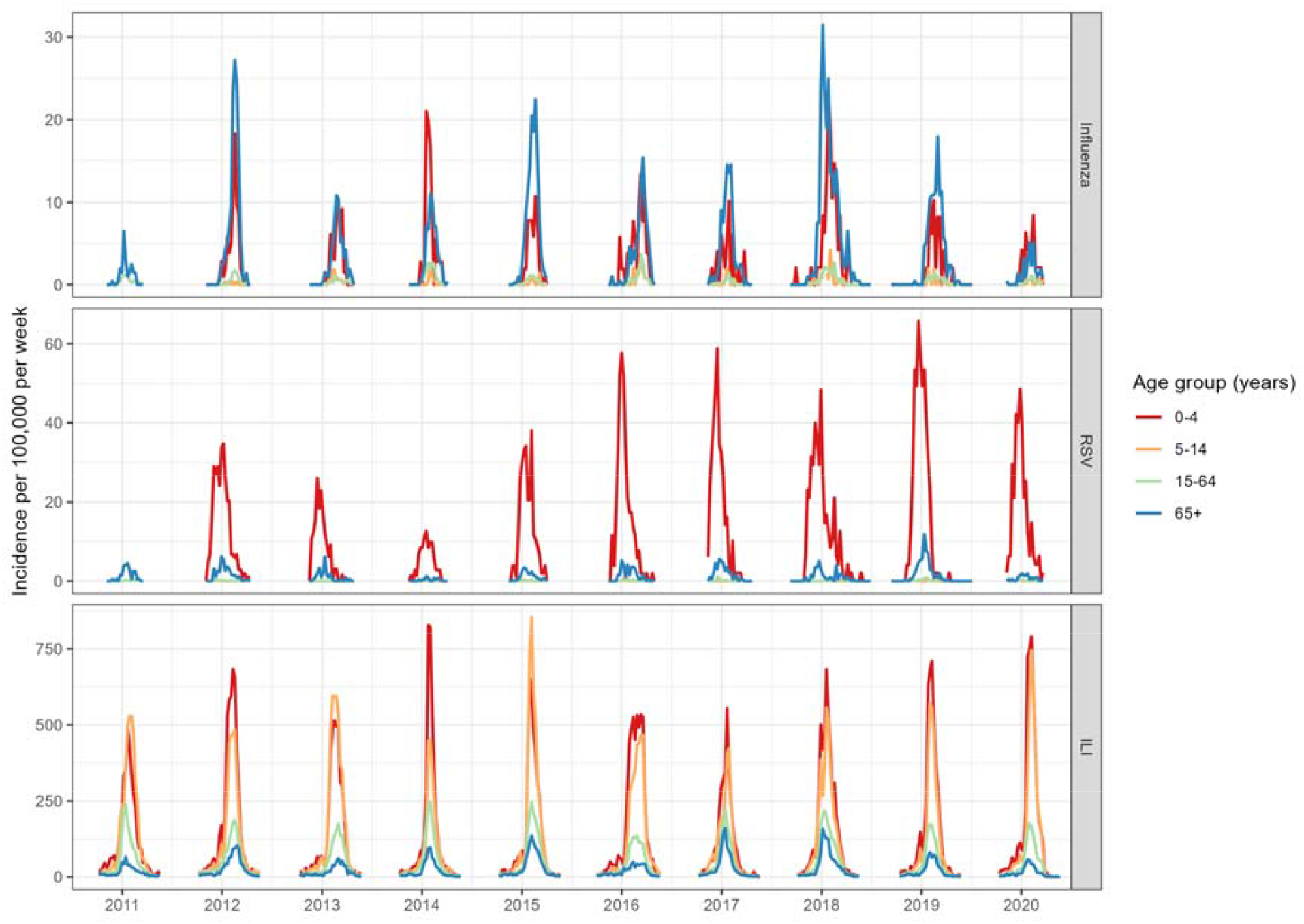
Weekly incidence of viral infections by age over 2010-2020. The figure shows the weekly hospitalisation incidence with influenza (top row) and respiratory syncytial virus (RSV) (middle row) reported by the Valencia Hospital Surveillance Network for the Study of Influenza and Other Respiratory Viruses (VAHNSI) study, and weekly rates of community influenza-like illness (ILI) per 100, 000 (bottom row) in sentinel sites in Spain over the period. Each colour represents an age group according to the wider age categories in the ILI dataset, with the smaller VAHNSI age categories aggregated to match these. Alt text: Line plots showing viral incidence over the study period, with line colours indicating the age groups, and the top, middle and bottom panels respectively showing influenza, respiratory syncytial virus and community influenza-like illness.

### Estimated baseline model with no interaction

The baseline model was fit to hospitalisation and ILI data simultaneously, estimating all non-interaction parameters (Figure 3 and Supplementary Table 2). The transmission rate was consistently higher in influenza (ranging from 1.9 to 2.8 transmissions per potentially infectious contact per week) compared with RSV (ranging from 1.2 to 2.0), and later introduction for influenza (mostly October and November) than for RSV (mostly August and September). Hospitalisation probabilities for both viruses were higher at very young and older ages. The probability of RSV hospitalisation per incident infection was far higher in younger age groups (6.4% in babies under 6 months compared with 0.2% in those 75 years or older), whereas the probability of influenza hospitalisation was more similar for these age groups (0.6% and 0.7% respectively, Figure 3). Children under 5 also had a higher probability of community consultation for ILI, but the probability was higher for an influenza infection (6.6%) compared with RSV (3.1%). For the oldest age group (65 and over), we did not observe an elevated probability of community consultation.

**Figure 3.**
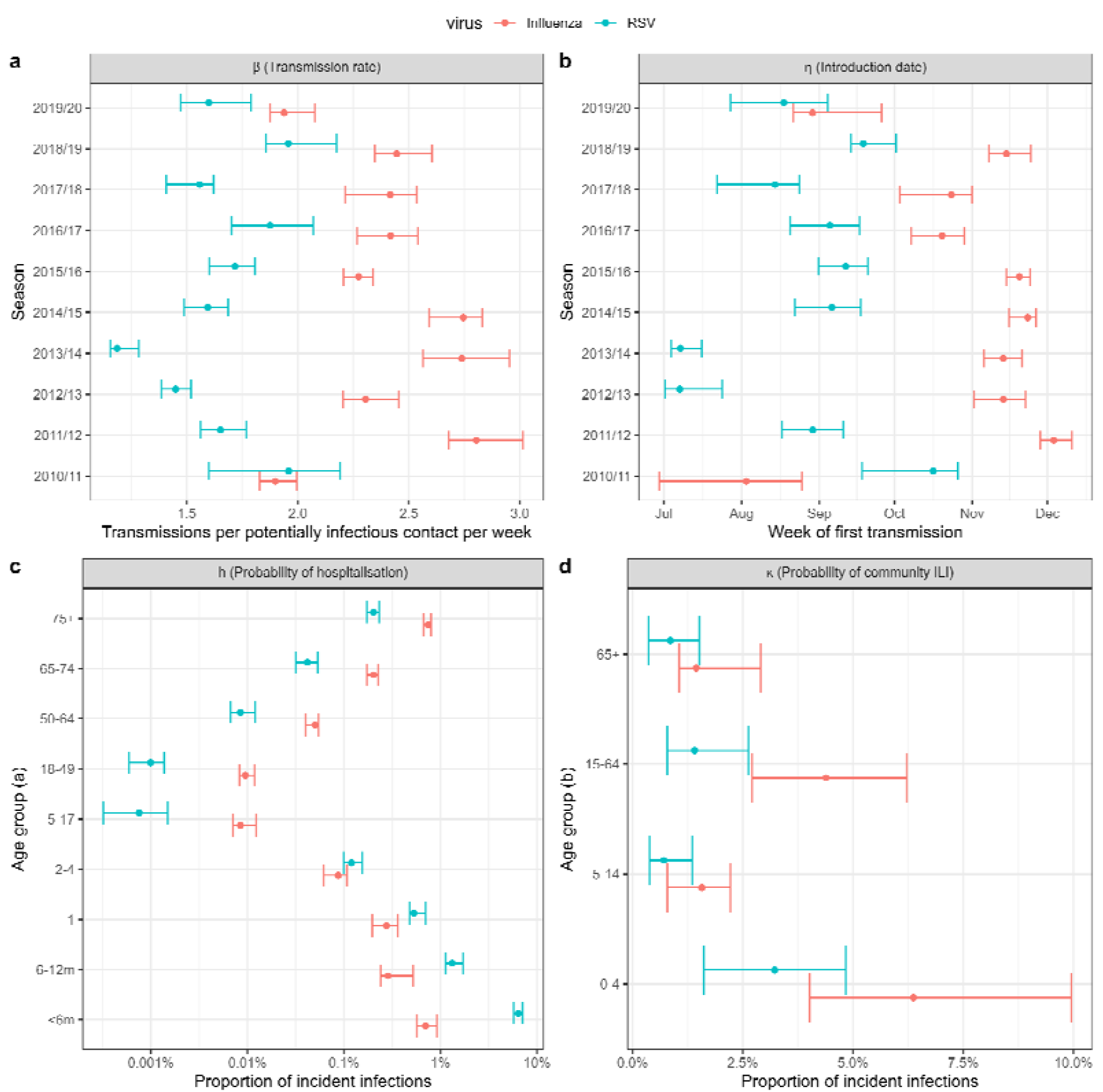
Estimates of non-interaction model parameters for influenza and RSV estimated in the baseline model. Parameters include: a) transmission rate (β) for each season (rows), b) the week of introduction (η) for each season (rows), c) the hospitalisation probability given incident infection (h) for each age-group (nine age categories, rows), and d) the probability of community consultation for ILI (κ) for each age group (four age categories, rows). The error bars show the 95% credibility interval from the posterior distribution. Alt text: Graphs with points and errors bars showing estimated model parameters, with different colours for each virus, and the four panels showing a) transmission rates, b) dates of introduction, c) hospitalisation probabilities and d) probabilities of community ILI.

### Estimated parameters characterising interaction

The interaction parameters were then independently estimated to assess each potential mechanism. Many interaction models improved the fit relative to the baseline, the best-fit single parameter model being one in which the hospitalisation probability of influenza was reduced to 20% of baseline during active infection with RSV (Table 1), with values between 0 and 60% improving the fit over the baseline model. The 2^nd^ best fitting single model included a 10-day refractory period following infection with influenza (range 2-17 days). Other interaction mechanisms which improved the fit included modification of the hospitalisation probability and the transmission rate of RSV during an influenza infection, and modification of susceptibility in both directions. Comparison with models where pairs of interaction parameters were allowed to vary (Supplementary Table 3) demonstrates that the best fitting paired models always include the same mechanism (active RSV reduces influenza severity, with similar effect size).

**Table 1.** Comparison of models with single interaction parameters. ΔAIC was calculated as the difference of the model-associated AIC with that of the baseline model. The parameter column designates the interaction parameter estimated in the fit. The estimated value for the best parameter set is that giving the highest likelihood, with the estimate interval in parentheses indicating the range of values which present an improvement over the baseline model, as estimated by a positive ΔAIC value. K refers to the total number of estimated parameters in each model. The full list of estimates for non-interaction parameters estimated for the baseline model is shown in Supplementary Table 2.

| Rank | Interaction parameter | Description | K | $\Delta AIC$ | Baseline | Estimate (improvement interval) |
| --- | --- | --- | --- | --- | --- | --- |
| Best-fit | $\theta_1$ | Hospitalisation with influenza affected by active infection with RSV | 67 | 52.5 | 1 | 0.2 (0-0.6) |
| 2<sup>nd</sup> best fit | $1/\psi_1$ | Duration of influenza refractory period $R_f$ (weeks) | 67 | 19.2 | 0,001 | 1.4 (0.3-2.4) |
| | $\sigma_2$ | Transmission of RSV affected by active infection with influenza | 67 | 18.3 | 1 | 0 (0-0.7) |
| | $\delta_2$ | Susceptibility to RSV affected by active infection with influenza | 67 | 10.0 | 1 | 0.4 (0-0.9) |
| | $\delta_1$ | Susceptibility to influenza affected by active infection with RSV | 67 | 4.7 | 1 | 0.9 (0.8-1) |
| | $\theta_2$ | Hospitalisation with RSV affected by active infection with influenza | 67 | 1.1 | 1 | 0.6 (0.3-0.8) |
| Baseline | - | No interaction | 66 | 0 |  |  |
| | $1/\omega_2$ | Duration of RSV residually positive period $R_v$ (weeks) | 67 | <0 | 0,001 | |
| | $1/\psi_2$ | Duration of RSV refractory period $R_f$ (weeks) | 67 | <0 | 0,001 | |
| | $\sigma_1$ | Transmission of influenza affected by active infection with RSV | 67 | <0 | 1 | |
| | $1/\omega_1$ | Duration of influenza residually positive period $R_v$ (weeks) | 67 | <0 | 0,001 | |

The single-interaction model which best fits the data captures much of the variation in peak height, and captures the few cases in which there were observed co-detections (Figure 4). Breakdowns of the model fits by age against the hospitalisation data (Supplementary Figure 2), and the fit against the community ILI data (Supplementary Figure 3) show a more variable quality of fit. Examination of the cumulative numbers of cases demonstrates that the interaction of active RSV infection with influenza hospitalisation probability had a negligible effect on RSV and influenza cases (0.0%), but a relatively large effect on the number of co-detections (63%, Figure 5). While the best-fitting model, with modified influenza hospitalisation probability, reduced co-detections in each season relative to the no interaction model, the second best-fitting model, with a refractory period following influenza infection, showed little difference from the no-interaction model in terms of co-detections (Supplementary Figure 4), suggesting that different mechanisms which improve the fit to data do so with relation to different aspects of the observed data.

**Figure 4.**
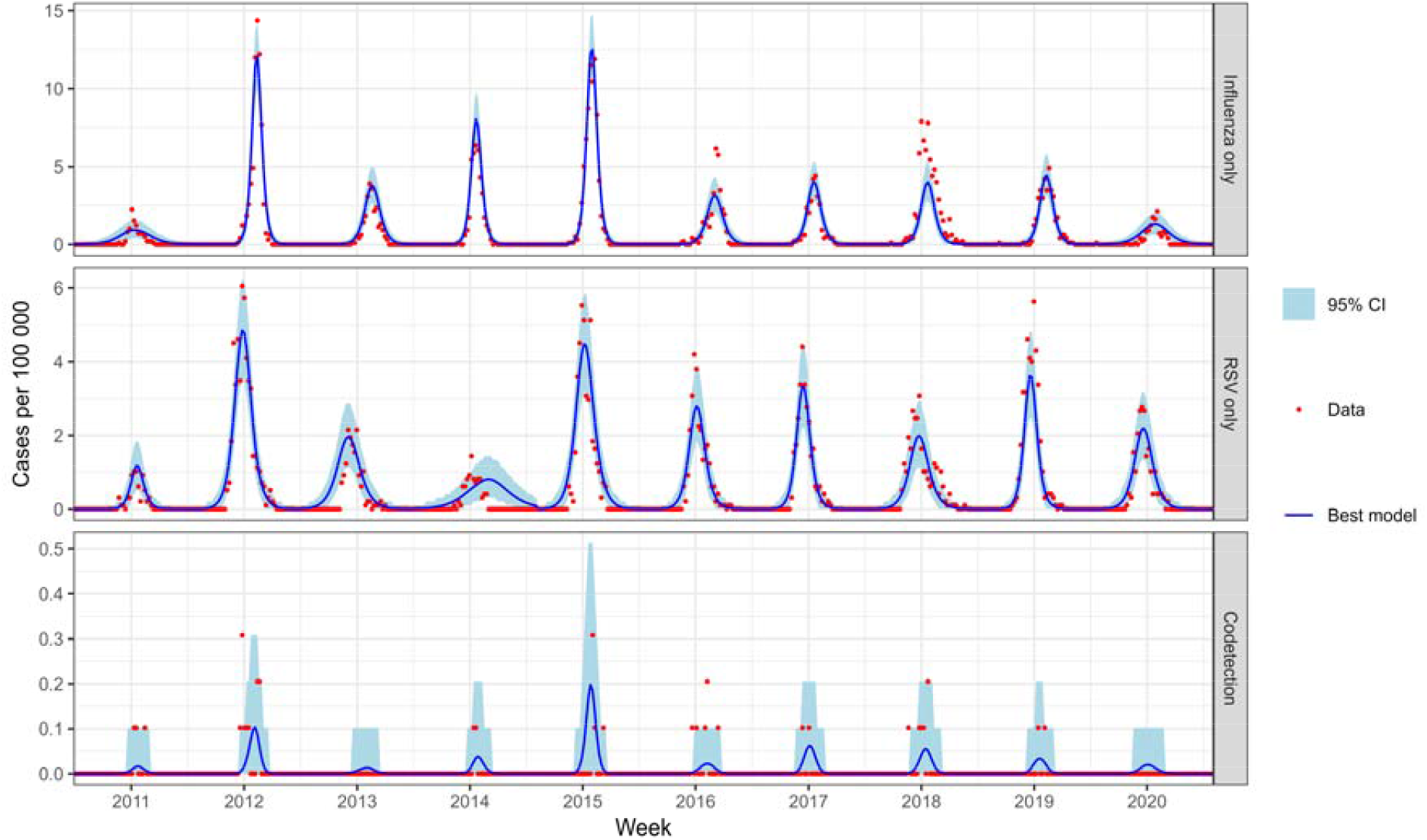
Number of cases of hospitalisation in the data and best fitting model (with variable) for each week. The top row indicates mono-detection of influenza, the middle row RSV mono-detection and the bottom row the co-detections of the two. The shaded area represents the 95% confidence interval of the model predictions based on a Poisson distribution. The data and model have been rescaled to represent hospitalisation incidence per 100,000 population. Alt text: Time series plots showing the best-fitting model (lines and shaded areas) against the data (points) over the study period, with the top, middle and bottom panels respectively showing influenza only, respiratory syncytial virus only and co-detection.

**Figure 5.**
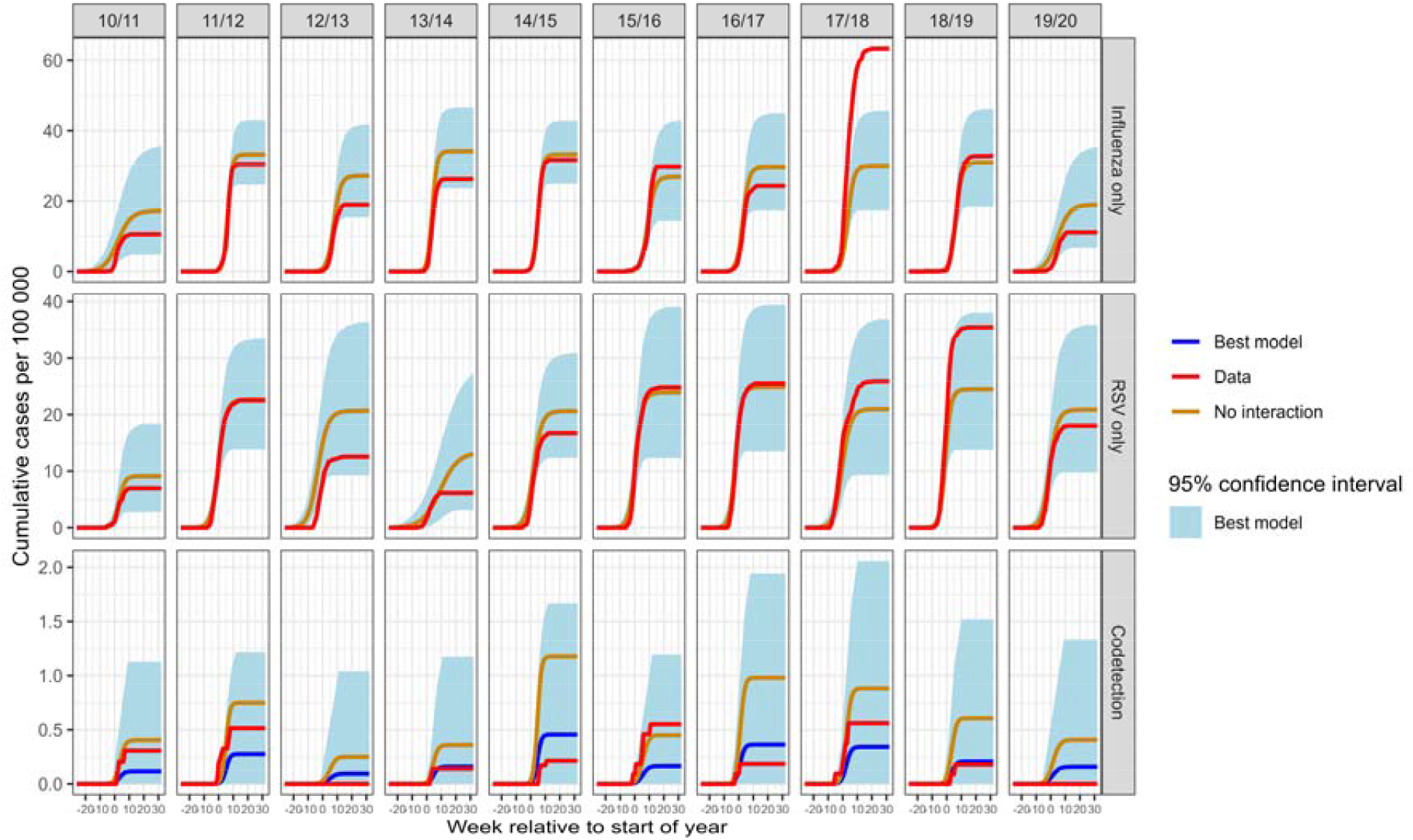
Cumulative number of cases of hospitalisation per 100,000 population under the best fitting model, the equivalent model with no interaction, and the data. The top row indicates mono-detection of influenza, the middle row RSV mono-detection and the bottom row the co-detections of the two. Each column represents a transmission season. The shaded area represents the 95% confidence interval of the model predictions based on a Poisson distribution. Alt text: Line plots showing the cumulative number of incident cases in each season, with colours indicating the best-fitting model, the non interaction model and the data. Each column is a season, while the top, middle and bottom rows respectively show influenza only, respiratory syncytial virus only and co-detection.

We also explored how the effect of the interaction would change if epidemics of influenza and RSV happened simultaneously (Figure 6). The hospitalisation rate (best) interaction had a considerable effect on the number of co-detections, with a similar relative effect even if the epidemics of occurred simultaneously. This interaction also had a negligible effect on overall case numbers. However, the refractory period (second best) interaction had very little effect on co-detections (as demonstrated in Supplementary Figure 4) but a much larger effect on numbers of RSV cases, especially with simultaneous epidemics.

**Figure 6.**
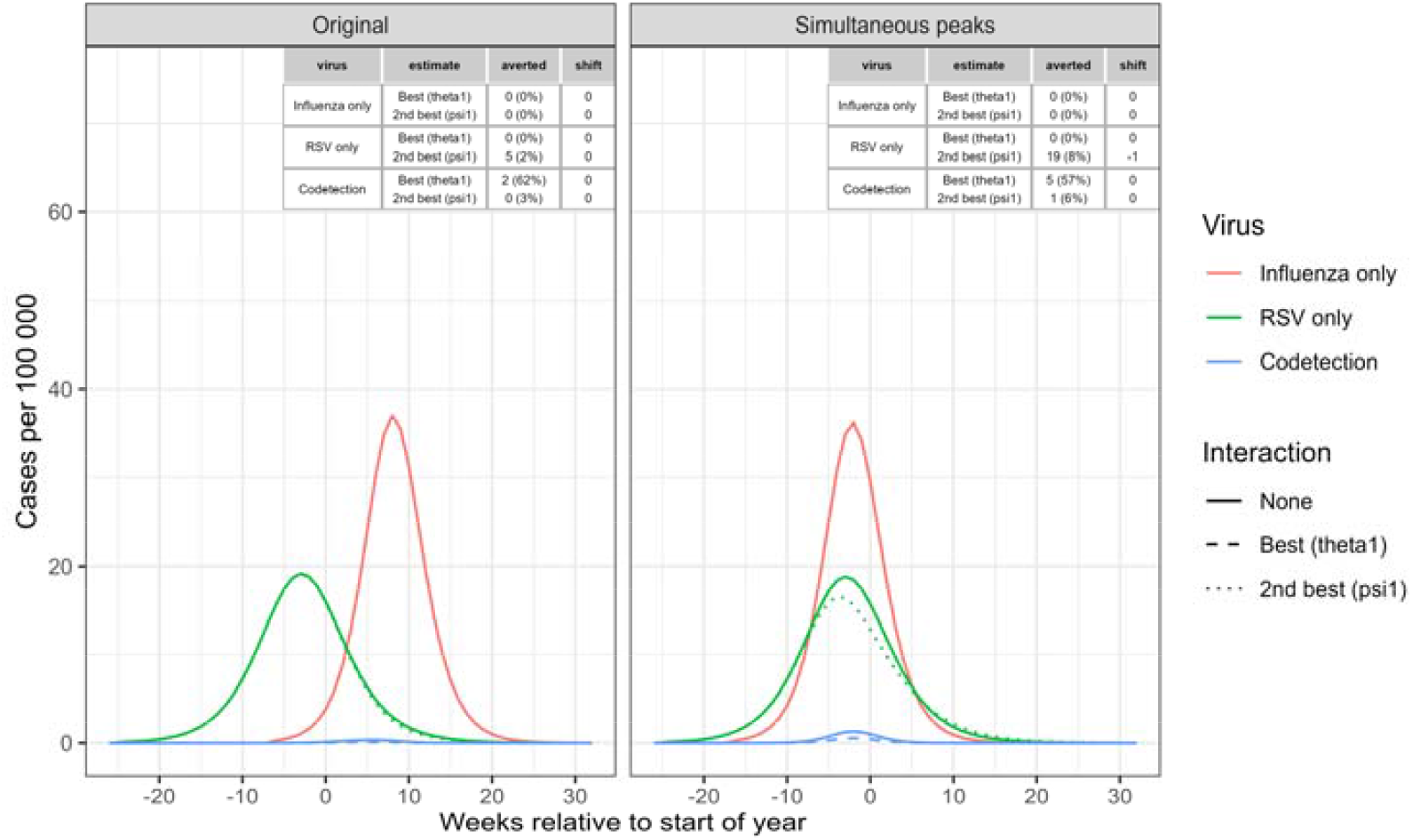
The effect of interaction on the number of cases of each virus and co-detections, in typical non-simultaneous epidemics (left panel) and epidemics modified to occur simultaneously (right panel). The line colour shows the modelled hospital incidence of each virus or co-detection, and the line type shows the level of interaction, with “None” representing the best fit model but with the interaction parameters set to neutral), and the best and second-best interaction model corresponding with the highest likelihood values from Table 1. The embedded table records the number and percentage of total cases averted as a result of the interaction, as well as the shift as a result of interaction in the week at which the peak occurs. Alt text: Line plot showing a transmission season, either with the original parameters (left panel), or with one epidemic shifted in time (right panel). The colours indicate the type of virus, the line types indicate the model, and an inset table in each panel shows the cases averted by viral interaction.

## Discussion

We propose a transmission model with two viruses to quantify interaction between influenza and RSV data, and apply this to a robust dataset from a 10-year active surveillance network in Valencia, Spain using multiplex molecular testing. We demonstrate that these data, which have few codetections, are best explained by a model including an interaction mechanism, which is best supported assuming that active RSV infection reduces the severity of incident influenza.

Our analysis additionally supports a model in which active RSV infection reduced influenza susceptibility, which is consistent with *in vitro work showin*g that RSV infection reduced cellular susceptibility to influenza A^7^. Our results also suggest possible effects of active influenza infection on RSV, including reduced transmission rate, reduced susceptibility and hospitalisation rate, as well as a refractory period following influenza infection in which the individual is protected from RSV. This is consistent with several other studies which have identified protective effects of influenza A infection on RSV ^8–10^. Interestingly, work in mice has demonstrated that prior infection with RSV reduced influenza A morbidity ^12^, but found no effect of influenza A on RSV ^12^, whereas the current analysis finds the inverse with respect to the refractory periods.

Population-level studies have pointed to lower than expected numbers of co-detections as evidence of inhibition between influenza and RSV ^4,17–21^. Several groups have used dynamic models to explain this apparent interaction. Surveillance on a 10-year cohort of young children found a possible period of cross-protection, consistent with our own analysis^36^. A model fit to 6 years of data from Canada and Hong Kong estimated the strength of viral interaction in reducing susceptibility, both during infection and for some time after recovery ^31^. The authors estimated a strong interaction signal in both sites, suggesting that RSV reduces subsequent susceptibility to influenza and vice versa, consistent with our own best estimates. Their estimated effect of influenza on RSV lasted over a timescale of months, whereas the effect of RSV on influenza lasted only weeks ^31^. Our analysis suggested a possible refractory period following influenza infection, but lasting only ten days, and does not support a refractory period following RSV infection. An age-structured model applied to weekly case counts from USA over 10+ years identified three possible interaction mechanisms by which might reproduce the data, namely by reducing susceptibility to RSV, reducing transmissibility of RSV (both of which are supported by results in our work), and increasing recovery rate from RSV (which we did not consider here) ^37^.

While several studies previously measured interactions between influenza and RSV at the population level, the dynamic model proposed here brings several advantages. Firstly, the use of an explicit observation process determining whether infections appear in our clinical datasets reduces collider bias present in typical association studies ^38^, whereby a criterion for selection into the sample, accessing medical treatment, is correlated with the presence of either virus, and therefore distorts the association between the two viruses. Secondly, the two-virus framework allows us to directly compare different interaction mechanisms operating from influenza to RSV and vice versa. In this capacity, we were able to evaluate a wide range of such mechanisms including a residual positivity stage which has not to our knowledge been previously considered. Thirdly, the data were collected over a 10-year period with consistent enrolment criteria throughout and systematic multiplex sampling, while also accounting for community ILI reporting rates. Data from molecular multiplex testing is increasingly available but its analysis remains challenging and requires explicit assumptions to link infection dynamics and observations, which this model provides.

A two-pathogen model, and a traditional age-structured compartmental model, each require multiple dimensions of compartments. In order to capture both facets simultaneously without requiring an encumbrant number of compartments, we developed an innovative method to model age structure. The outbreak in each season was modelled as a single simultaneous epidemic across all age groups, but which was then observed in the age-specific data according to the relative contact rate of each age group, and an age-specific reporting rate. While the timings of outbreak between different age groups might contribute to low numbers of co-detections, previous analysis does not indicate major temporal heterogeneity^19^.

To compensate the necessary complexity of a two-pathogen model, several simplifying assumptions were made, introducing potential limitations. Firstly, due to small numbers of cases, we did not consider different subtypes of influenza or RSV, treating each as a single outbreak in each season.

This implies that subtypes had the same epidemiological characteristics (e.g. transmission rate, interactions) and perfect cross-protection among strains. Test accuracy was not accounted for, assuming that false positive rates were negligible (with the exception of residual positivity), and allowing for the false negative rate to be subsumed in the reporting rates. We also did not account for the contribution of other respiratory viruses to community ILI, due to the dominance of these two viruses as causes of ILI^39^.

Secondly, we did not account for seasonality of transmission. Instead, we considered seasons were independent and estimated the date of the introduction in each season. Temperature and humidity are known drivers of influenza transmission ^40^, and consequently some studies model evolving transmission rates ^31^. Although these assumptions could impact our estimates related to single virus transmissibility, they should not affect our results regarding interactions.

Thirdly, we did not include super-seasonal or waning immunity, preferring to assume that all individuals were susceptible at the start of each season but then fully protected for the rest of the season after infection. Age-specific differences in immunity have been implicitly captured by the participation probability and reporting rates.

Lastly, we assumed that hospitalisation can only occur at a single point in disease progression, that risk of hospitalisation varied only with respect to age, while in reality hospitalisation depends on many patient characteristics and may happen at multiple points over the course of infection^41,42^.

Additionally, hospitalised cases were not removed from the pool of transmitters, although because the proportion of hospitalised cases were rare (less than 8% even for the highest risk age group), this is considered negligible.

The rollout of preventive treatments for RSV including monoclonal antibodies for newborns, and vaccines for elderly people and pregnant women to target newborns, may substantially affect RSV incidence. Specific studies should be designed to examine the impact of interaction under new immunisation and vaccination schedules and campaigns. Recent evidence suggests that influenza vaccination reduces incidence of RSV hospitalisation in young children ^43^, but further work should examine bi-directional effects. Fundamental studies should further explore the biological mechanisms of these interactions in natural infections. Cohort studies in different age groups should investigate immune correlations of co-infection or sequential infection. Further modelling work should explore interactions with other viruses such as metapneumovirus, rhinovirus and SARS-CoV-2, especially in light of potential new combination vaccines ^44^. Interestingly, the 2025-2026 season in Europe was characterized by early and high influenza circulation and a delayed RSV epidemic ^45^.

Whether this was caused by the expansion of new preventive measures against RSV remains to be determined.

To conclude, our work supports the existence of inhibitory interactions between influenza and RSV. Such interactions should be considered when evaluating the introduction of new vaccination and immunization campaigns and their timing. A strong inhibitory interaction could mitigate the effect of a vaccination campaign, or cause a shift in epidemic timing with unexpected effects on preparations for seasonal illness.

## Author contributions

Conceptualization: G.S., S.S.C., L.C., F.X.L.L., L.O.

Data curation: B.M.C., A.M.I., J.P.B., A.O.S., J.D.D.

Formal analysis: G.S.

Funding acquisition: S.S.C., L.C., F.X.L.L., L.O., B.M.C., A.M.I., J.P.B., A.O.S., J.D.D.

Investigation: G.S., L.O.

Methodology: G.S., L.O.

Project administration: S.S.C., L.C., F.X.L. L., L.O.

Resources: L.C., F.X.L.L.

Software: G.S. Supervision: L.O.

Validation: G.S. Visualization: G.S.

Writing – original draft: G.S., L.O.

Writing – review & editing: G.S., S.S.C., L. C., F.X.L.L., L.O.

## Valencia Hospital Surveillance Network for the Study of Influenza and Other Respiratory Viruses (VAHNSI)

Additional members of the VAHNSI group include: Laura Cano Pérez, Mario Carballido Fernández, Juan Mollar Maseres, Miguel Tortajada Girbés, Germán Schwarz Chávarri, Vicente Gil Guillén, Ramón Limón Ramírez, Empar Carbonell Franco, Angel Belenguer Varea, Concepción Carratalá Munuera, José Vicente Tuells Hernández.

## Ethical approval

The Ethics Research Committee of the Dirección General de Salud Pública-Centro Superior de Investigación en Salud Pública (DGSP-CSISP) approved the original protocol of the study, described in ^25^, and our analysis was based on this data in aggregate.

## Code and data availability

The code used to conduct the analysis and produce the figures is available from github.com/georgeshirreff/ViralInteraction, along with the national surveillance data and a subset of the multiplex data. The complete anonymised and aggregated multiplex data on which the study is based are available on reasonable request from the corresponding author. The data are not publicly available due to privacy restrictions.

