## Supplementary Information for "Interactions between influenza and respiratory syncytial viruses: two-pathogen modelling to analyse multiplex data"

### Supplementary methods

##### Sampling procedure

The data collection period spanned from 2010-2021 across 11 participating hospitals. The denominator population was the total estimated catchment population for each age group across all hospitals enrolled within each season (Supplementary Table 4). The sampling was generally conducted during the putative influenza seasons (November to March inclusive) between mid-2010 and mid-2021, with the exceptions of 2017/18 and 2018/19 when the sampling season was deliberately extended for the purposes of the study (September to June in 2017/18, September to August in 2018/2019), and in 2020 when sampling was disrupted by the COVID-19 pandemic (September to early March in 2019/20, and December to May in 2020/21). Specific dates of study for each season are shown in Supplementary Table 4. Children under 18 were not included in 2010/11 but were included thereafter. Data from the 2020/21 season was excluded as this was disrupted by the COVID-19 pandemic and therefore not representative.

Potential participants were those admitted to participating hospitals with acute respiratory illness. Dedicated nurses screened all hospitalised patients discharged from the Emergency Department if the referral was possibly related to a respiratory infection. To qualify for enrolment in the study, patients needed to be resident in the catchment area of one of the participating hospitals, non-institutionalised, to have been admitted <48 hours before enrolment and not have been previously discharged from a hospital within 30 days prior to the current admission. Only patients with acute respiratory illness, having symptom onset <7 days before hospital admission date, were considered for enrolment. The symptoms required for enrolment were age specific. Patients ≥5 years of age had to fit the European Centre for Disease Prevention and Control clinical case definition of influenza-like illness (ILI), meaning they had at least one systemic symptom (fever or feverishness, headache, myalgia, or malaise) and one respiratory symptom (cough, sore throat, or shortness of breath). For children <5 years, the inclusion criteria was broad, including any clinical conditions potentially associated with a presentation of acute respiratory illness (list of conditions reported in Supplementary Table 5). Clinical and demographic characteristics from patients were obtained by a face-to-face interview or by consulting medical records. Informed consent was taken before enrolment from patients, or their legal guardians where appropriate ^25^.

Enrolled patients had oropharyngeal and nasopharyngeal swabs collected if aged ≥14 years, or nasal and nasopharyngeal swabs if <14 years. Each patient was sampled at a single time point. Both swabs were combined in one tube of viral transport media (Copan, Italy) and frozen at or below -20°C at the study site until shipped refrigerated to the coordinating site’s centralized virology laboratory ^25^.

##### Laboratory analysis

One third of the viral transport medium volume (1 mL) was used for total nucleic acids extraction using an automated silica-based method (Nuclisens Easy-Mag, BioMérieux, Lyon, France). Subsequently, extracted nucleic acids were analysed using a real time multiplex reverse transcription polymerase chain reaction (RT-PCR) panel, testing for the presence of Respiratory Syncytial Virus (A/B, RSV) and influenza viruses (A/B) ^26,27^ as described previously ^25^. From the start of the study to the 2013/14 season, the master-mix AgPath-ID™ One-Step RT-PCR Kit (Ambion, USA) was used, while from 2014/15 onwards the master-mix QScript XLT One-Step RT-qPCR ToughMix (Quantabio, MA, USA) was used. From 2014/15 onwards the primers and probes were also updated for RSV (to include new RSV-B circulating clades) ^46^. An external laboratory was used to conduct the analyses in the 2010/11 and 2011/12 seasons. A co-detection sample is defined as one that is positive for more than one viral species.

##### Aggregation

The hospitalisation dataset used was aggregated by week and across nine age groups (<6 months; 6-12 months; 1 year; 2-4 years; 5-17 years; 18-49 years; 50-64 years; 65-74 years; and ≥75 years), as described in previous analysis ^19^. National incidence rates of ILI were accessed by week across four age groups (≤4 years, 5-14 years, 15-64 years, ≥65 years). Week numbers were defined as in ^47^ counting from the final week of the calendar year, week 0, and weeks prior to this are negative numbers.

##### Model parameters

Parameters are defined according to virus *v* (1 for influenza, 2 for RSV), age group *a* with nine categories, and season *s* (2010/11 to 2019/20 inclusive). An additional age group category *b* represented the age distribution of community ILI data, with only four age categories (Supplementary Table 4). Sources of parameter values are shown in Supplementary Table 1.

##### Age-specific contact rates

Average contact rates by age group were estimated from the POLYMOD survey data ^32,48^ using the R package *socialmixr* ^32^. As there was no specific data for Spain, the entire dataset, representing data from Belgium, Germany, Finland, the United Kingdom, Italy, Luxembourg, the Netherlands, and Poland, was used. The participants were separated into age groups and the total number of contacts (physical and non-physical) per participant was averaged across the age group. As the POLYMOD data did not distinguish partial years, the <12m contacts were assumed to apply to both the <6m and 6-12m age groups. These were standardised relative to the highest contact age group 5-17 years to give the age-specific relative probability of participating in the epidemic, $\pi_{a}$.

#### Model state equations

The total population size N, which remains constant, is the sum over the population size of all compartments:

$N=\sum_{X\in S,E,I,R,Rv,Rf,R} \sum_{Y\in S,E,I,R,Rv,Rf,R} XY$

Progression from E to I occurred at virus-specific progression rate $\alpha_{v}$, from I to Rv at rate $\gamma_{v}$, from Rv to Rf at rate $\omega_{v}$ and from Rf to R at rate $\psi_{v}$ (Supplementary Table 1). The following differential equations determine the rates of movement of individuals between internal compartments, sub-headed by the state with respect to virus 1.

##### S with respect to virus 1

$$\frac{dSS}{dt}=-\left( \lambda_{1}+\lambda_{2} \right)SS$$

$$\frac{dSE}{dt}=\lambda_{2}SS-\left( \lambda_{1}+\alpha_{2} \right)SE$$

$$\frac{dSI}{dt}=\alpha_{2}SE-\left( \lambda_{1}\delta_{1}+\gamma_{2} \right)SI$$

$$\frac{dSRv}{dt}=\gamma_{2}SI-\left( \lambda_{1}+\omega_{2} \right)SRv$$

$$\frac{dSRf}{dt}=\omega_{2}SRv-\left( \lambda_{1}0+\psi_{2} \right)SRf$$

$$\frac{dSR}{dt}=\psi_{2}SRf-\lambda_{1}SR$$

##### E with respect to virus 1

$$\frac{dES}{dt}=\lambda_{1}SS-\left( \alpha_{1}+\lambda_{2} \right)ES$$

$$\frac{dEE}{dt}=\lambda_{1}SE+\lambda_{2}ES-\left( \alpha_{1}+\alpha_{2} \right)EE$$

$$\frac{dEI}{dt}=\lambda_{1}\delta_{1}SI+\alpha_{2}EE-\left( \alpha_{1}+\gamma_{2} \right)EI$$

$$\frac{dERv}{dt}=\lambda_{1}SRv+\gamma_{2}EI-\left( \alpha_{1}+\omega_{2} \right)ERv$$

$$\frac{dERf}{dt}=\lambda_{1}0SRf+\omega_{2}ERv-\left( \alpha_{1}+\psi_{2} \right)ERf$$

$$\frac{dER}{dt}=\lambda_{1}SR+\psi_{2}ERf-\alpha_{1}ER$$

##### I with respect to virus 1

$$\frac{dIS}{dt}=\alpha_{1}ES-\left( \gamma_{1}+\lambda_{2}\delta_{2} \right)IS$$

$$\frac{dIE}{dt}=\alpha_{1}EE+\lambda_{2}\delta_{2}IS-\left( \gamma_{1}+\alpha_{2} \right)IE$$

$$\frac{dII}{dt}=\alpha_{1}EI+\alpha_{2}IE-\left( \gamma_{1}+\gamma_{2} \right)II$$

$$\frac{dIRv}{dt}=\alpha_{1}ERv+\gamma_{2}II-\left( \gamma_{1}+\omega_{2} \right)IRv$$

$$\frac{dIRf}{dt}=\alpha_{1}ERf+\omega_{2}IRv-\left( \gamma_{1}+\psi_{2} \right)IRf$$

$$\frac{dIR}{dt}=\alpha_{1}ER+\psi_{2}IRf-\gamma_{1}IR$$

##### Rv with respect to virus 1

$$\frac{dRvS}{dt}=\gamma_{1}IS-\left( \omega_{1}+\lambda_{2} \right)Rv$$

$$\frac{dRvE}{dt}=\gamma_{1}IE+\lambda_{2}RvS-\left( \omega_{1}+\alpha_{2} \right)Rv$$

$$\frac{dRvI}{dt}=\gamma_{1}II+\alpha_{2}RvE-\left( \omega_{1}+\gamma_{2} \right)Rv$$

$$\frac{dRvRv}{dt}=\gamma_{1}IRv+\gamma_{2}RvI-\left( \omega_{1}+\omega_{2} \right)Rv$$

$$\frac{dRvRf}{dt}=\gamma_{1}IRf+\omega_{2}RvRv-\left( \omega_{1}+\psi_{2} \right)Rv$$

$$\frac{dRvR}{dt}=\gamma_{1}IR+\psi_{2}RvRf-\omega_{1}Rv$$

##### Rf with respect to virus 1

$$\frac{dRfS}{dt}=\omega_{1}RvS-\left( \psi_{1}+\lambda_{2}0 \right)RfS$$

$$\frac{dRfE}{dt}=\omega_{1}RvE+\lambda_{2}0RfS-\left( \psi_{1}+\alpha_{2} \right)RfE$$

$$\frac{dRfI}{dt}=\omega_{1}RvI+\alpha_{2}RfE-\left( \psi_{1}+\gamma_{2} \right)RfI$$

$$\frac{dRfRv}{dt}=\omega_{1}RvRv+\gamma_{2}RfI-\left( \psi_{1}+\omega_{2} \right)RfRv$$

$$\frac{dRfRf}{dt}=\omega_{1}RvRf+\omega_{2}RfRv-\left( \psi_{1}+\psi_{2} \right)RfRf$$

$$\frac{dRfR}{dt}=\omega_{1}RvR+\psi_{2}RfRf-\psi_{1}RfR$$

##### R with respect to virus 1

$$\frac{dRS}{dt}=\psi_{1}RfS-\lambda_{2}RS$$

$$\frac{dRE}{dt}=\psi_{1}RfE+\lambda_{2}RS-\alpha_{2}RE$$

$$\frac{dRI}{dt}=\psi_{1}RfI+\alpha_{2}RE-\gamma_{2}RI$$

$$\frac{dRRv}{dt}=\psi_{1}RfRv+\gamma_{2}RI-\omega_{2}RRv$$

$$\frac{dRRf}{dt}=\psi_{1}RfRf+\omega_{2}RRv-\psi_{2}RRf$$

$$\frac{dRR}{dt}=\psi_{1}RfR+\psi_{2}RRf$$

The transmission of infection to susceptible individuals is determined by the forces of infection for each virus. The force of infection for each virus is calculated by multiplying the transmission rate $\beta_{vs}$ by the proportion of individuals infectious with virus *v* (with modification by the modified infectiousness parameter $\sigma_{v}$ if appropriate):

$$\lambda_{1}=\left( Import\left( t,\eta_{1} \right)+\beta_{1}\left( IS+IE+II\sigma_{1}+IRv+IRf+IR \right) \right)N^{-1}$$

$$\lambda_{2}=\left( Import\left( t,\eta_{2} \right)+\beta_{2}\left( SI+EI+II\sigma_{2}+RvI+RfI+RI \right) \right)N^{-1}$$

There is additionally an importation process which introduces $\rho$ new infections over a 1.5 week period spanning time $\eta_{vs}$. The importation rate is determined by a trapezoid step function in which the total number of imported cases for each virus is $\rho$, and these are introduced over a 1.5 week time period centred on time $\eta_{vs}$ for each virus *v* and season *s*. The importation rate at a particular time *t* given importation time $\eta_{vs}$is as follows, and this is also displayed graphically in Supplementary Figure 5. The 1.5 week time period was chosen such that the area under the curve sums to $\rho$.

$$Import\left( t,\eta_{v} \right)=\rho\cdot\left\{ \begin{matrix} 0 & \text{if} t<\eta_{v}-0.75 \\ \left( t-\left( \eta_{v}-0.75 \right) \right)/0.5 & \text{if} \eta_{v}-0.75< t<\eta_{v}-0.25 \\ 1 & \text{if} \eta_{v}-0.25<t<\eta_{v}+0.25 \\ \left( t - \left( \eta_{v}+ 0.75 \right) \right)/-0.5 & \text{if}\eta_{v}+0.25<t<\eta_{v}+0.75 \\ 0 & \mathrm{if} \eta_{v}+0.75<t \end{matrix} \right.$$

#### Model observation equations

During a single numerical integration, the epidemic of both viruses is simulated for each season and virus. This describes the timing and size of the epidemic in the age group with the highest contact rate. We calculate the incidence of infections (not necessarily seeking treatment either by hospitalisation or by community consultation for ILI) of each virus on a given day and whether or not they also would test positive for the second virus (2+ or 2-) as follows:

$$\mathrm{Inc}_{1,2+}\left( t \right)=\int_{t-1}^{t} \alpha_{1}\left( EI\left( t \right)+ERv\left( t \right) \right)$$

$$\mathrm{Inc}_{2,1+}\left( t \right)=\int_{t-1}^{t} \alpha_{2}\left( IE\left( t \right)+RvE\left( t \right) \right)$$

$$\mathrm{Inc}_{1,2\text{-}}\left( t \right)=\int_{t-1}^{t} \alpha_{1}\left( ES\left( t \right)+EE\left( t \right)+ERf\left( t \right)+ER\left( t \right) \right)$$

$$\mathrm{Inc}_{2,1\text{-}}\left( t \right)=\int_{t-1}^{t} \alpha_{2}\left( SE\left( t \right)+EE\left( t \right)+ RfE\left( t \right)+RE\left( t \right) \right)$$

The outbreak was assumed to occur simultaneously across all age groups, but with different age-related participation probabilities $\pi_{a}$ referring to their relative probability of becoming infected, compared to the highest contact age group. Of those incident infections, a proportion *h_va_* were recorded as hospitalised cases. The probability of hospitalisation because of incident virus 1 was modified during active infection with virus 2 by the parameter $\theta_{1}$, and vice versa by parameter $\theta_{2}$. The total number of hospitalisations testing positive for virus 1, virus 2 and for both viruses together in age group *a* was calculated as follows:

$$\mathrm{Hosp}_{1only,a}\left( t \right)=\left( \mathrm{Inc}_{1,2\text{-}}\left( t \right)+\mathrm{Inc}_{1,2\text{+}}\left( t \right)\theta_{1}\left( 1-\pi_{a} \right) \right)\pi_{a}h_{1a}$$

$$\mathrm{Hosp}_{2only,a}\left( t \right)=\left( \mathrm{Inc}_{2,1\text{-}}\left( t \right)+\mathrm{Inc}_{2,1\text{+}}\left( t \right)\theta_{2}\left( 1-\pi_{a} \right) \right)\pi_{a}h_{2a}$$

$$\mathrm{Hosp}_{12,a}\left( t \right)=\left( \mathrm{Inc}_{1,2\text{+}}\left( t \right)h_{1a}\theta_{1}+\mathrm{Inc}_{2,1\text{+}}\left( t \right){h_{2a}\theta}_{2} \right)\pi_{a}\pi_{a}$$

In addition to the incidence of cases arriving through hospitalisation, we also modelled the weekly rates of reported community ILI by a sentinel network, assuming that incident infections were reported as ILI at a rate $\kappa_{vb}$ for each virus *v* and age group *b*. Reporting ILI was assumed to be independent of hospitalisation. As the age categories were larger for the ILI data than those used in the model, these were aggregated to the larger age categories by multiplying the total number of incident cases of ILI by the participation probabilities $\pi_{a}$ weighted by the relative population size of age group *a* in age group *b*, where $P_{a}$ is the size of the catchment population for age group *a*. This total number of reported ILI cases was finally divided by the total modelled population size *N* to give the overall per capita ILI rate. As the boundaries in the ILI data did not exactly match those in the hospital data, we assumed that the rate of ILI in the 5-14 age group was similar to that of the 5-17 age group, and likewise for the 15-64 and the 18-64 age groups, and aggregated the groups accordingly.

$${ILI rate}_{b}\left( t \right)=\left( \left( \mathrm{Inc}_{1,2\text{-}}\left( t \right)+\mathrm{Inc}_{1,2\text{+}}\left( t \right) \right)\kappa_{1b}+ \left( \mathrm{Inc}_{2,1\text{-}}\left( t \right)+\mathrm{Inc}_{2,1\text{+}}\left( t \right) \right)\kappa_{2b} \right)\frac{\sum_{a \epsilon b} P_{a}\pi_{a}}{\sum_{a \epsilon b} P_{a}}N^{-1}$$

#### Likelihood equations

The likelihood of observing the number of hospitalised cases in each week and age group given the model was calculated for each virus individually and their co-detections, assuming that these followed a Poisson distribution. The likelihood of community ILI for each week and age group was calculated assuming a log-normal distribution, and assuming that no other viruses besides influenza and RSV contributed to ILI incidence.

*Hospital-based reporting.* We assumed that hospitalisations, given as numbers of cases per week in each age group *a* of mono-detections for each virus, $Y_{1only,a}\left( t \right)$ and $Y_{2only,a}\left( t \right)$, and co-detection data $Y_{12,a}\left( t \right)$, followed a Poisson distribution. A Poisson log-likelihood with parameter the modelled hospitalisations was therefore considered. The expression of the log-likelihood is given below:

$\mathcal{L}\left( Y|\chi\right)=\sum_{a} \sum_{t} \left[ \begin{aligned} \ln\left( Pois\left( Y_{1only,a}\left( t \right)|\mathrm{Hosp}_{1only,a}\left( t \right)\frac{{Pop}_{at}}{N} \right) \right)+ \\ \ln\left( Pois\left( Y_{2only,a}\left( t \right)|\mathrm{Hosp}_{2only,a}\left( t \right)\frac{{Pop}_{at}}{N} \right) \right)+ \\ \ln\left( Pois\left( Y_{12,a}\left( t \right)|\mathrm{Hosp}_{12,a}\left( t \right)\frac{{Pop}_{at}}{N} \right) \right) \end{aligned} \right]$

where $Pois\left( Y|\lambda\right)$ is the Poisson density function for value $Y$ given mean $\lambda$:

$$Pois\left( Y|\lambda\right)=\frac{\lambda^{Y}e^{-\lambda}}{Y!}$$

*Community-based reporting.* ILI incidence in the community was analysed as a rate per week *t* per age group *b* per 100,000 people and denoted $W_{b}\left( t \right)$. The likelihood of observing the data given the model was calculated according to a log-normal distribution, as the ILI data were continuous and highly dispersed. The mean was given by the log of the modelled value and the standard deviation by the standard deviation of the log values of all the community ILI data values, $W$. For both the calculation of the standard deviation and the calculation of likelihood, 1 was added to both the observed and modelled values.

$$\mathcal{L}\left( ILI|\chi\right)=\sum_{b}^{\begin{aligned} 0\text{-}4,5\text{-}14, \\ 15\text{-}64,65+ \end{aligned}} \sum_{t} \ln\left( LogNorm\left( W_{b}\left( t \right)+1 | \text{ln}\left( {ILI rate}_{b}\left( t \right)+1 \right), sdLogW \right) \right)$$

where *sd* is the standard deviation of the log data:

$$sd=\sqrt{\frac{\sum\left( \ln\left( W+1 \right)-\frac{\sum\ln\left( W+1 \right)}{n} \right)^{2}}{n-1}}$$

and $LogNorm\left( Y|m,sd \right)$ is the lognormal density function for a value Y given log mean *m* and log standard deviation *s*:

$$LogNorm\left( Y|m,sd \right)=\frac{1}{Ysd\sqrt{2\pi}}e^{-\frac{1}{2}\left( \frac{\ln\left( Y \right)-m}{sd} \right)^{2}}$$

#### Model inference process

Parameters were estimated using an adaptive Markov Chain Monte Carlo (MCMC) approach with a Metropolis Hastings algorithm, *modMCMC* from the *FME* package^35^.

First, the baseline model was estimated, assuming no interaction, meaning that all interaction parameters were fixed to baseline values, while the non-interaction parameters (transmission rate, season start time, hospitalisation probability and community ILI consultation probability) were estimated. As each season was considered to be independent, with parameter values specific only to that season, parameter proposals were conducted using a blocked Gibbs approach to allow for more efficient convergence ^49^. In Iteration 1, new values were proposed for all variable parameters, then for all non-seasonal parameters for the second iteration. In the following 10 subsequent iterations, only parameters specific to each season were proposed, before the cycle began again (a schematic of the process of model inference is provided in Supplementary Figure 6). This was run for 100,000 iterations with a burn-in of 20,000 iterations, repeating the inference of each model 6 times from different starting points to ensure convergence. The parameter values which produced the highest likelihood were recorded as $\hat{\chi_{v}}$. The best AIC value was calculated for the baseline model (K=66 parameters).

We then fit models in which non-interaction parameters were fixed, and a single interaction parameter was allowed to vary, to assess where this improved the fit to the data. Model ability to improve on the baseline model was based on calculation of the AIC. For each interaction model with a single varying interaction parameter, the AIC was calculated for each posterior parameter value (using K=66+1). An interaction model was considered to have improved the fit if any AIC value was smaller than that of the baseline AIC. For any model providing an improvement, the most likely parameter value was reported, as well as the range of values which corresponded to an AIC smaller than the baseline model. Parameter value estimates were not provided for models which did not improve the fit.

### Supplementary Figures

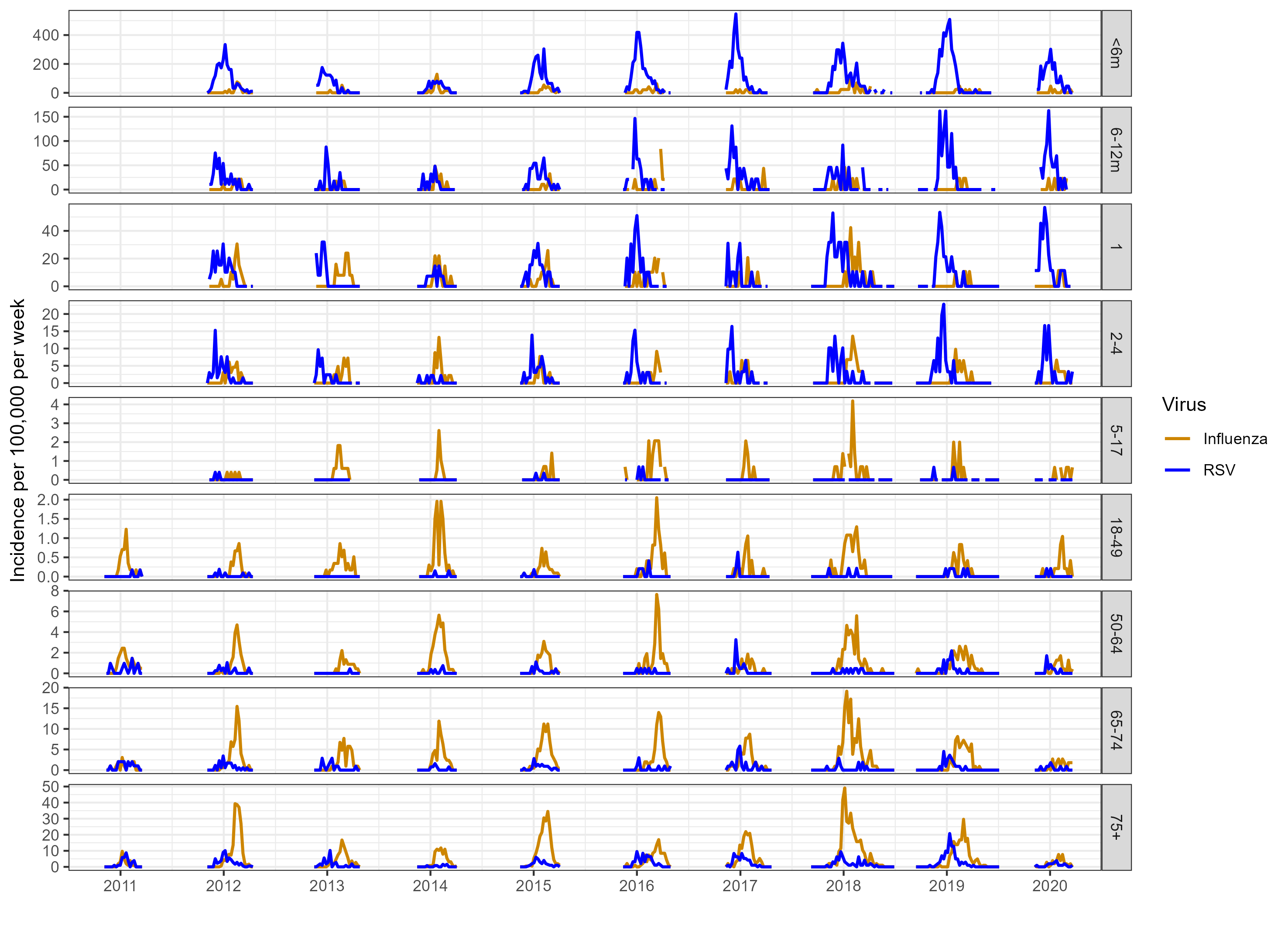

Supplementary Figure 1. Weekly incidence per 100,000 individuals of influenza and RSV infections by age over 2010-2020 as reported by the VAHNSI surveillance network study. Each row represents an age group, and the colours represents the two viruses.

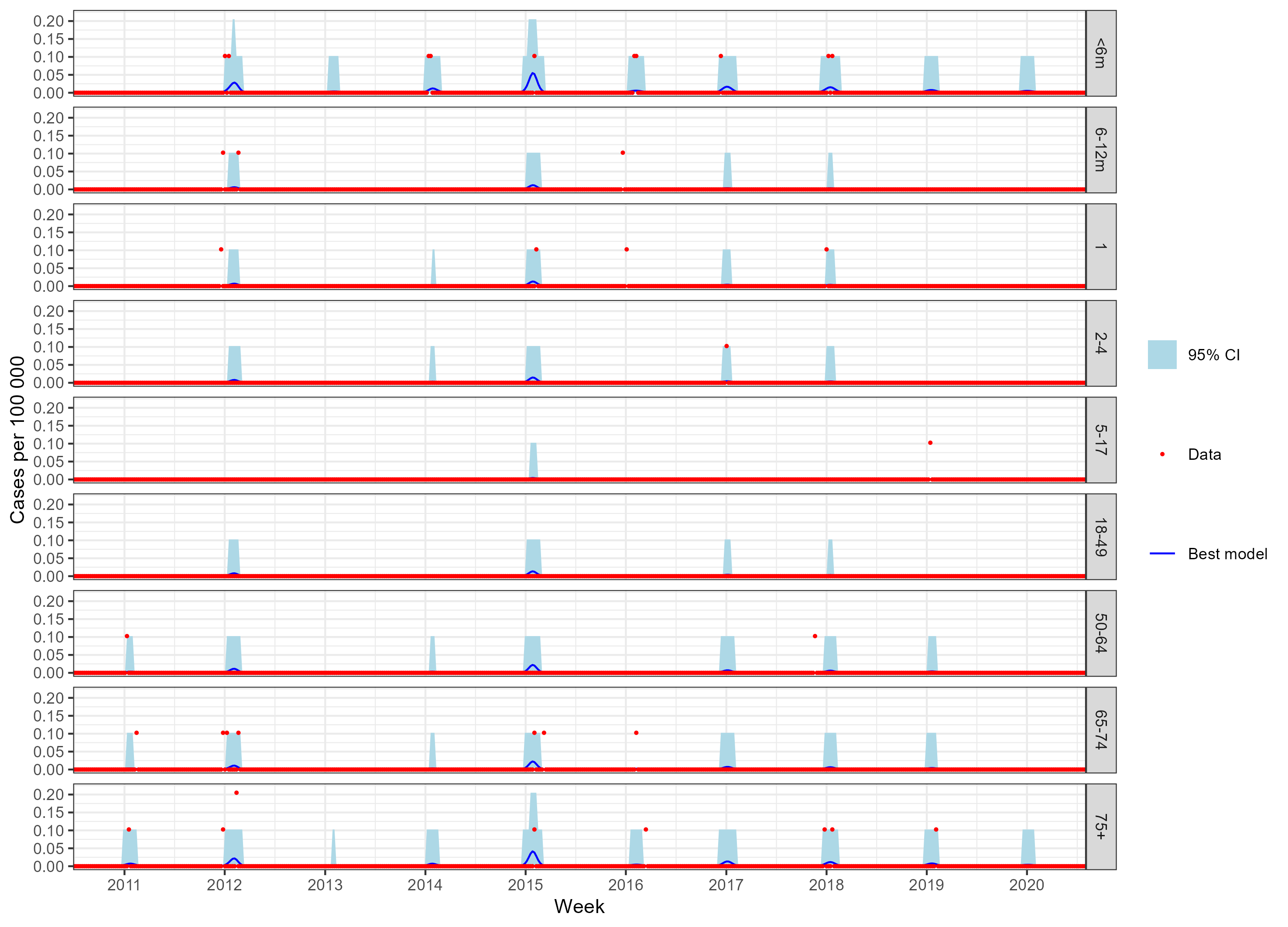

Supplementary Figure 2. Model fit of influenza-RSV co-detection by season and age-groups. Number of co-detection cases in the VAHNSI hospitalisation data (red) and best fitting model (blue, with variable $\delta_{1}$, representing a 60% reduction in susceptibility to influenza during active infection with RSV) for each week, with each row representing a different age group. The grey area represents the 95% confidence interval of the model based on a Poisson distribution.

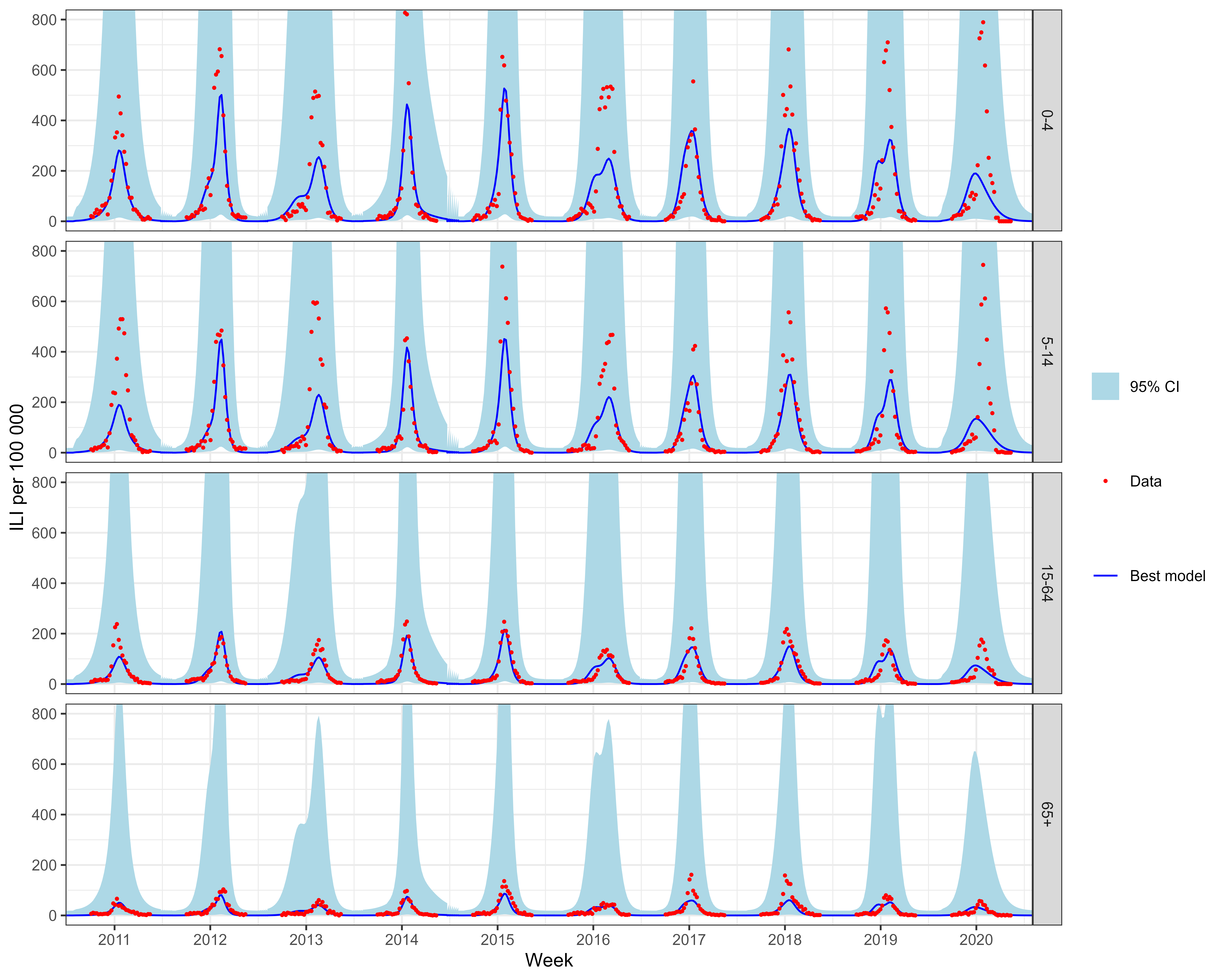

Supplementary Figure 3. Model fits of community ILI by season and age groups. Weekly ILI incidence rate per 100 000 individuals from sentinel sites in Spain (red) and corresponding predictions (blue, with variable $\delta_{1}$, representing a 60% reduction in susceptibility to influenza during active infection with RSV). The rows represent the different age groups. The light blue area represents the 95% confidence interval of the model predictions based on a log-normal distribution.

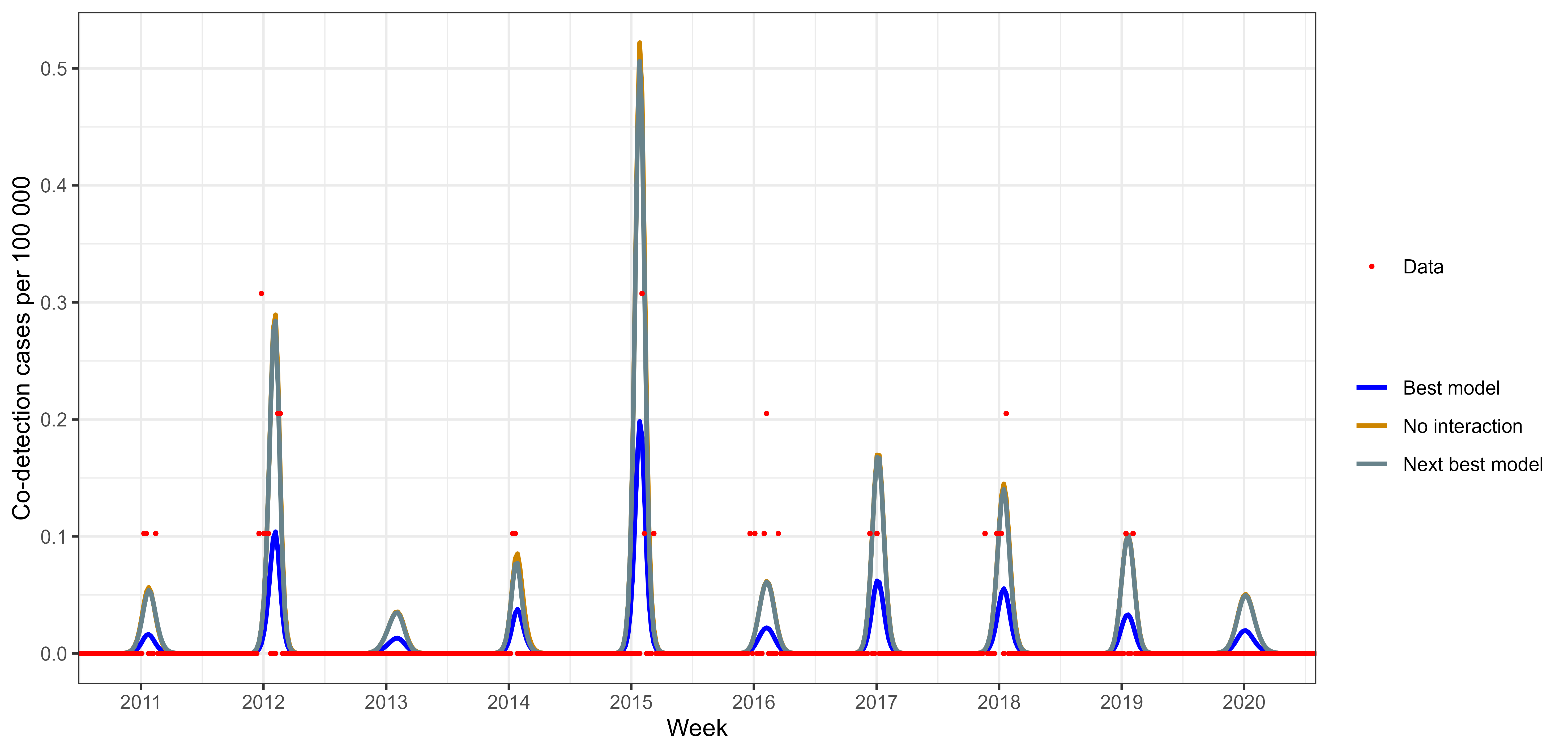

Supplementary Figure 4. Model fit to weekly co-detections in hospitalization data showing model comparison, over all seasons. Co-detection cases per 100,000 individuals in the data and across the baseline (brown, no interaction), best fitting model (blue, with variable $\delta_{1}$, representing a 60% reduction in susceptibility to influenza during active infection with RSV) and 2^nd^ best fitting model (grey, with variable $\theta_{1}$, representing an 80% reduction in probability of hospitalisation with influenza during active infection with RSV). Cases are shown by week, with the columns representing the season.

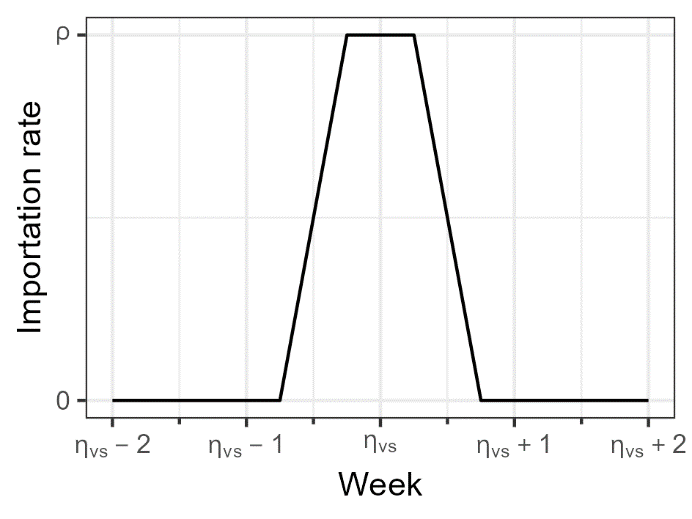

Supplementary Figure 5. Function showing the current importation rate changing as a proportion of the maximum rate $\rho$ at each week relative to the modelled date of introduction $\eta_{vs}$.

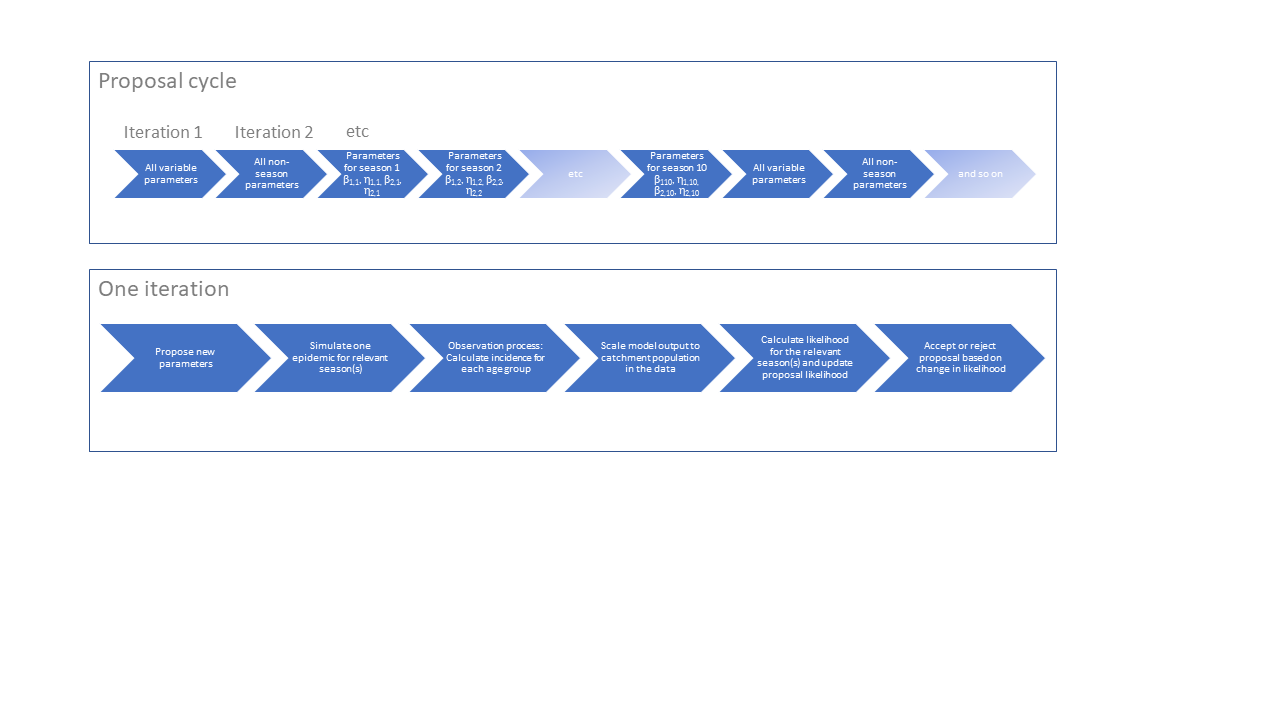

Supplementary Figure 6. Schematic showing the process of statistical inference. Top) the cycle of proposals for the Markov Chain Monte Carlo estimation, where each arrow represents one iteration, and the text indicates for which parameters new values are proposed. The pale arrows represent the pattern continuing for several steps. Bottom) The modelling process required for a single iteration of the MCMC, with each arrow representing a single step in the process.

### Supplementary Tables

Supplementary Table 1. Description of model parameters.

| Category | Parameter | Symbol | Value | Source |
| --- | --- | --- | --- | --- |
|  | Transmission rate (rate of infection per week) | $\beta_{vs}$ | By virus, season | Estimated (range 0,5) |
| Non-interaction | Date of introduction (week) | $\eta_{vs}$ | By virus, season | Estimated (range -26,25) |
|  | Hospitalisation probability | $h_{va}$ | By virus, age group | Estimated (range 10^-8^,1) |
|  | Community ILI consultation probability | $\kappa_{vb}$ | By virus, age group | Estimated (range 0,1) |
|  | Population size, initial susceptible population | $N$ | Hospital catchment population | Supplementary Table 4 |
|  | Importations (per week for each virus) | $\rho$ | 2000 | Arbitrary |
|  | Probability of participation in the epidemic | $\pi_{a}$ | By age group | Derived from POLYMOD age-specific contact rates^32^ |
| Fixed | Duration of latency (weeks) | $\frac{1}{\alpha_{v}}$ | Influenza: 2.61 days | ^50^* |
|  |  |  | RSV: 4.5 days |  |
|  | Duration of active infection (weeks) | $\frac{1}{\gamma_{v}}$ | Influenza: 4.58 days | ^50^ |
|  |  |  | RSV: 7.72 days |  |
| Interaction | Modification of susceptibility | $\delta_{v}$ | By virus pair | Estimated (baseline 1) |
|  | Modification of infectiousness | $\sigma_{v}$ | By virus pair | Estimated (baseline 1) |
|  | Modification of hospitalisation probability | $\theta_{v}$ | By virus pair | Estimated (baseline 1) |
|  | Duration of residually positive period (weeks) | $\frac{1}{\omega_{v}}$ | By virus pair | Estimated (baseline 0.001 weeks) |
|  | Duration of refractory period (weeks) | $\frac{1}{\psi_{v}}$ | By virus pair | Estimated (baseline 0.001 weeks) |
|  | *The source paper reports incubation period, which we have used as a proxy for latency period | | | |

Supplementary Table 2. Best estimates of all parameters in the best-fitting baseline model. The estimates represent those in the highest likelihood parameter set (mode), with the 95% credibility interval from the posterior distribution using a χ-squared likelihood comparison test relative to the highest likelihood.

| Parameter | Virus | Age group | Season | Estimate |
| --- | --- | --- | --- | --- |
| $\beta$ | Influenza | all | 2010/11 | 1.9 (1.83-2) |
|  |  |  | 2011/12 | 2.8 (2.68-3.01) |
|  |  |  | 2012/13 | 2.31 (2.2-2.45) |
|  |  |  | 2013/14 | 2.74 (2.56-2.95) |
|  |  |  | 2014/15 | 2.75 (2.59-2.83) |
|  |  |  | 2015/16 | 2.27 (2.21-2.34) |
|  |  |  | 2016/17 | 2.42 (2.27-2.54) |
|  |  |  | 2017/18 | 2.42 (2.21-2.54) |
|  |  |  | 2018/19 | 2.45 (2.35-2.6) |
|  |  |  | 2019/20 | 1.94 (1.88-2.08) |
|  | RSV |  | 2010/11 | 1.96 (1.6-2.19) |
|  |  |  | 2011/12 | 1.65 (1.56-1.77) |
|  |  |  | 2012/13 | 1.45 (1.39-1.52) |
|  |  |  | 2013/14 | 1.19 (1.16-1.28) |
|  |  |  | 2014/15 | 1.59 (1.49-1.69) |
|  |  |  | 2015/16 | 1.72 (1.6-1.81) |
|  |  |  | 2016/17 | 1.88 (1.7-2.07) |
|  |  |  | 2017/18 | 1.56 (1.41-1.62) |
|  |  |  | 2018/19 | 1.96 (1.86-2.18) |
|  |  |  | 2019/20 | 1.6 (1.47-1.79) |
| $\eta$ | Influenza |  | 2010/11 | Jul 29 (Jun 25-Aug 21) |
|  |  |  | 2011/12 | Nov 28 (Nov 23-Dec 05) |
|  |  |  | 2012/13 | Nov 06 (Oct 25-Nov 15) |
|  |  |  | 2013/14 | Nov 05 (Oct 28-Nov 12) |
|  |  |  | 2014/15 | Nov 14 (Nov 06-Nov 17) |
|  |  |  | 2015/16 | Nov 16 (Nov 11-Nov 21) |
|  |  |  | 2016/17 | Oct 14 (Oct 02-Oct 23) |
|  |  |  | 2017/18 | Oct 17 (Sep 27-Oct 25) |
|  |  |  | 2018/19 | Nov 07 (Oct 31-Nov 17) |
|  |  |  | 2019/20 | Aug 21 (Aug 13-Sep 17) |
|  | RSV |  | 2010/11 | Oct 12 (Sep 14-Oct 22) |
|  |  |  | 2011/12 | Aug 24 (Aug 12-Sep 05) |
|  |  |  | 2012/13 | Jun 30 (Jun 24-Jul 17) |
|  |  |  | 2013/14 | Jun 29 (Jun 25-Jul 08) |
|  |  |  | 2014/15 | Aug 28 (Aug 13-Sep 08) |
|  |  |  | 2015/16 | Sep 08 (Aug 28-Sep 17) |
|  |  |  | 2016/17 | Aug 31 (Aug 15-Sep 12) |
|  |  |  | 2017/18 | Aug 08 (Jul 16-Aug 18) |
|  |  |  | 2018/19 | Sep 11 (Sep 06-Sep 24) |
|  |  |  | 2019/20 | Aug 09 (Jul 19-Aug 27) |
| $h$ | Influenza | <6m | common | 0.701% (0.567%-0.918%) |
|  |  | 6-12m |  | 0.286% (0.24%-0.52%) |
|  |  | 1 |  | 0.275% (0.197%-0.362%) |
|  |  | 2-4 |  | 0.0871% (0.0616%-0.107%) |
|  |  | 5-17 |  | 0.00846% (0.00707%-0.0123%) |
|  |  | 18-49 |  | 0.00952% (0.00827%-0.0119%) |
|  |  | 50-64 |  | 0.0502% (0.0401%-0.0544%) |
|  |  | 65-74 |  | 0.202% (0.173%-0.226%) |
|  |  | 75+ |  | 0.744% (0.669%-0.798%) |
|  | RSV | <6m |  | 6.38% (5.73%-7.08%) |
|  |  | 6-12m |  | 1.32% (1.13%-1.71%) |
|  |  | 1 |  | 0.53% (0.477%-0.701%) |
|  |  | 2-4 |  | 0.12% (0.0994%-0.154%) |
|  |  | 5-17 |  | 0.000756% (0.000322%-0.00149%) |
|  |  | 18-49 |  | 0.000994% (0.000594%-0.00138%) |
|  |  | 50-64 |  | 0.00846% (0.00668%-0.012%) |
|  |  | 65-74 |  | 0.0418% (0.0318%-0.0534%) |
|  |  | 75+ |  | 0.202% (0.172%-0.232%) |
| $\kappa$ | Influenza | 0-4 | common | 6.37% (4.01%-9.95%) |
|  |  | 5-17 |  | 1.57% (0.788%-2.22%) |
|  |  | 15-64 |  | 4.39% (2.71%-6.22%) |
|  |  | 65+ |  | 1.44% (1.06%-2.91%) |
|  | RSV | 0-4 |  | 3.22% (1.62%-4.84%) |
|  |  | 5-17 |  | 0.71% (0.386%-1.36%) |
|  |  | 15-64 |  | 1.41% (0.787%-2.63%) |
|  |  | 65+ |  | 0.858% (0.362%-1.51%) |

Supplementary Table 3. Interaction models comparisons with one or two parameters. ΔAIC was calculated as the difference of the model-associated AIC with that of the baseline model. The estimated value for the best parameter set is that giving the highest likelihood, with the estimation intervals in parentheses indicating the range of values which present an improvement over the baseline model, as estimated by a positive ΔAIC value. K is the total number of estimated parameters.

| Param1 | Description1 | Baseline1 | Param2 | Description2 | Baseline2 | K | ΔAIC | Estimate1 | Estimate2 |
| --- | --- | --- | --- | --- | --- | --- | --- | --- | --- |
| $\theta_{1}$ | Hospitalisation with influenza affected by active infection with RSV | 1 | ${1/\psi}_{1}$ | Duration of influenza refractory period Rf (weeks) | 0,001 | 68 | 68.5 | 0.2 (0.1-0.6) | 1.3 (0-2.4) |
| $\theta_{1}$ | Hospitalisation with influenza affected by active infection with RSV | 1 | $\sigma_{2}$ | Transmission of RSV affected by active infection with influenza | 1 | 68 | 66.3 | 0.2 (0.1-0.6) | 0 (0-1.1) |
| $\theta_{1}$ | Hospitalisation with influenza affected by active infection with RSV | 1 |  |  |  | 67 | 52.5 | 0.2 (0-0.6) |  |
| $\theta_{1}$ | Hospitalisation with influenza affected by active infection with RSV | 1 | $\delta_{2}$ | Susceptibility to RSV affected by active infection with influenza | 1 | 68 | 51.6 | 0.2 (0-0.7) | 0.8 (0.3-1.3) |
| $\theta_{1}$ | Hospitalisation with influenza affected by active infection with RSV | 1 | ${1/\omega}_{1}$ | Duration of influenza residually positive period Rv (weeks) | 0,001 | 68 | 51.3 | 0.2 (0-0.5) | 0.2 (0-1) |
| $\theta_{1}$ | Hospitalisation with influenza affected by active infection with RSV | 1 | $\delta_{1}$ | Susceptibility to influenza affected by active infection with RSV | 1 | 68 | 51.1 | 0.2 (0-0.8) | 1 (0.9-1.1) |
| $\theta_{1}$ | Hospitalisation with influenza affected by active infection with RSV | 1 | $\theta_{2}$ | Hospitalisation with RSV affected by active infection with influenza | 1 | 68 | 51.0 | 0.2 (0-0.5) | 1.2 (0.4-2.3) |
| $\theta_{1}$ | Hospitalisation with influenza affected by active infection with RSV | 1 | $\sigma_{1}$ | Transmission of influenza affected by active infection with RSV | 1 | 68 | 51.0 | 0.2 (0-0.6) | 1 (0.9-1.1) |
| $\theta_{1}$ | Hospitalisation with influenza affected by active infection with RSV | 1 | ${1/\psi}_{2}$ | Duration of RSV refractory period Rf (weeks) | 0,001 | 68 | 50.6 | 0.2 (0-0.5) | 0 (0-0.1) |
| $\theta_{1}$ | Hospitalisation with influenza affected by active infection with RSV | 1 | ${1/\omega}_{2}$ | Duration of RSV residually positive period Rv (weeks) | 0,001 | 68 | 50.5 | 0.2 (0-0.8) | 0 (0-0.3) |
| $\delta_{1}$ | Susceptibility to influenza affected by active infection with RSV | 1 | $\sigma_{1}$ | Transmission of influenza affected by active infection with RSV | 1 | 68 | 47.8 | 0.4 (0.2-0.8) | 2 (1.3-2.6) |
| ${1/\psi}_{1}$ | Duration of influenza refractory period Rf (weeks) | 0,001 | $\delta_{1}$ | Susceptibility to influenza affected by active infection with RSV | 1 | 68 | 23.4 | 1.4 (0-2.3) | 0.9 (0.8-1) |
| $\delta_{1}$ | Susceptibility to influenza affected by active infection with RSV | 1 | $\sigma_{2}$ | Transmission of RSV affected by active infection with influenza | 1 | 68 | 21.5 | 0.9 (0.8-1) | 0 (0-0.9) |
| ${1/\psi}_{1}$ | Duration of influenza refractory period Rf (weeks) | 0,001 | $\sigma_{2}$ | Transmission of RSV affected by active infection with influenza | 1 | 68 | 21.0 | 0.6 (0-2.7) | 0 (0-2.7) |
| $\delta_{2}$ | Susceptibility to RSV affected by active infection with influenza | 1 | $\sigma_{2}$ | Transmission of RSV affected by active infection with influenza | 1 | 68 | 21.0 | 0.6 (0-1.3) | 0 (0-1) |
| ${1/\psi}_{1}$ | Duration of influenza refractory period Rf (weeks) | 0,001 | $\delta_{2}$ | Susceptibility to RSV affected by active infection with influenza | 1 | 68 | 19.7 | 1.1 (0-2.2) | 0.7 (0.1-1.3) |
| $\theta_{2}$ | Hospitalisation with RSV affected by active infection with influenza | 1 | ${1/\psi}_{1}$ | Duration of influenza refractory period Rf (weeks) | 0,001 | 68 | 19.5 | 0.6 (0.1-1.5) | 1.3 (0.5-2.6) |
| ${1/\psi}_{1}$ | Duration of influenza refractory period Rf (weeks) | 0,001 |  |  |  | 67 | 19.2 | 1.4 (0.3-2.4) |  |
| $\theta_{2}$ | Hospitalisation with RSV affected by active infection with influenza | 1 | $\sigma_{2}$ | Transmission of RSV affected by active infection with influenza | 1 | 68 | 18.6 | 0.6 (0-1.6) | 0 (0-0.7) |
| $\sigma_{2}$ | Transmission of RSV affected by active infection with influenza | 1 |  |  |  | 67 | 18.3 | 0 (0-0.7) |  |
| ${1/\psi}_{1}$ | Duration of influenza refractory period Rf (weeks) | 0,001 | ${1/\psi}_{2}$ | Duration of RSV refractory period Rf (weeks) | 0,001 | 68 | 17.2 | 1.4 (0.2-2.7) | 0 (0-0.1) |
| ${1/\psi}_{1}$ | Duration of influenza refractory period Rf (weeks) | 0,001 | $\sigma_{1}$ | Transmission of influenza affected by active infection with RSV | 1 | 68 | 17.2 | 1.3 (0.4-2.2) | 1 (0.9-1.1) |
| ${1/\omega}_{2}$ | Duration of RSV residually positive period Rv (weeks) | 0,001 | $\sigma_{2}$ | Transmission of RSV affected by active infection with influenza | 1 | 68 | 16.4 | 0 (0-0.1) | 0 (0-0.8) |
| $\sigma_{1}$ | Transmission of influenza affected by active infection with RSV | 1 | $\sigma_{2}$ | Transmission of RSV affected by active infection with influenza | 1 | 68 | 16.4 | 1 (0.9-1.1) | 0 (0-0.7) |
| ${1/\psi}_{2}$ | Duration of RSV refractory period Rf (weeks) | 0,001 | $\sigma_{2}$ | Transmission of RSV affected by active infection with influenza | 1 | 68 | 16.4 | 0 (0-0.1) | 0 (0-0.7) |
| ${1/\omega}_{1}$ | Duration of influenza residually positive period Rv (weeks) | 0,001 | $\sigma_{2}$ | Transmission of RSV affected by active infection with influenza | 1 | 68 | 16.4 | 0 (0-0.3) | 0 (0-0.8) |
| $\theta_{2}$ | Hospitalisation with RSV affected by active infection with influenza | 1 | $\delta_{2}$ | Susceptibility to RSV affected by active infection with influenza | 1 | 68 | 14.5 | 9.7 (0.4-10) | 0.1 (0-0.9) |
| ${1/\omega}_{1}$ | Duration of influenza residually positive period Rv (weeks) | 0,001 | $\delta_{2}$ | Susceptibility to RSV affected by active infection with influenza | 1 | 68 | 13.9 | 0.5 (0-1.2) | 0 (0-0.8) |
| $\delta_{1}$ | Susceptibility to influenza affected by active infection with RSV | 1 | $\delta_{2}$ | Susceptibility to RSV affected by active infection with influenza | 1 | 68 | 12.3 | 0.9 (0.8-1) | 0.5 (0-1) |
| $\delta_{2}$ | Susceptibility to RSV affected by active infection with influenza | 1 |  |  |  | 67 | 10.0 | 0.4 (0-0.9) |  |
| $\sigma_{1}$ | Transmission of influenza affected by active infection with RSV | 1 | $\delta_{2}$ | Susceptibility to RSV affected by active infection with influenza | 1 | 68 | 8.1 | 1 (0.9-1.1) | 0.4 (0-0.8) |
| ${1/\omega}_{2}$ | Duration of RSV residually positive period Rv (weeks) | 0,001 | $\delta_{2}$ | Susceptibility to RSV affected by active infection with influenza | 1 | 68 | 8.1 | 0 (0-0.1) | 0.4 (0-0.9) |
| ${1/\psi}_{2}$ | Duration of RSV refractory period Rf (weeks) | 0,001 | $\delta_{2}$ | Susceptibility to RSV affected by active infection with influenza | 1 | 68 | 8.1 | 0 (0-0.1) | 0.4 (0-0.8) |
| $\theta_{2}$ | Hospitalisation with RSV affected by active infection with influenza | 1 | $\delta_{1}$ | Susceptibility to influenza affected by active infection with RSV | 1 | 68 | 4.7 | 0.6 (0.2-1.2) | 0.9 (0.8-1) |
| $\delta_{1}$ | Susceptibility to influenza affected by active infection with RSV | 1 |  |  |  | 67 | 4.7 | 0.9 (0.8-1) |  |
| $\delta_{1}$ | Susceptibility to influenza affected by active infection with RSV | 1 | ${1/\psi}_{2}$ | Duration of RSV refractory period Rf (weeks) | 0,001 | 68 | 2.8 | 0.9 (0.9-1) | 0 (0-0) |
| $\delta_{1}$ | Susceptibility to influenza affected by active infection with RSV | 1 | ${1/\omega}_{2}$ | Duration of RSV residually positive period Rv (weeks) | 0,001 | 68 | 2.7 | 0.9 (0.9-1) | 0 (0-0) |
| ${1/\omega}_{1}$ | Duration of influenza residually positive period Rv (weeks) | 0,001 | $\delta_{1}$ | Susceptibility to influenza affected by active infection with RSV | 1 | 68 | 2.7 | 0 (0-0.1) | 0.9 (0.9-1) |
| $\theta_{2}$ | Hospitalisation with RSV affected by active infection with influenza | 1 |  |  |  | 67 | 1.1 | 0.6 (0.3-0.8) |  |
| BASELINE | no interaction |  |  |  |  | 66 | 0 |  |  |
| $\theta_{2}$ | Hospitalisation with RSV affected by active infection with influenza | 1 | ${1/\omega}_{1}$ | Duration of influenza residually positive period Rv (weeks) | 0,001 | 68 | <0 |  |  |
| $\theta_{2}$ | Hospitalisation with RSV affected by active infection with influenza | 1 | ${1/\omega}_{2}$ | Duration of RSV residually positive period Rv (weeks) | 0,001 | 68 | <0 |  |  |
| $\theta_{2}$ | Hospitalisation with RSV affected by active infection with influenza | 1 | $\sigma_{1}$ | Transmission of influenza affected by active infection with RSV | 1 | 68 | <0 |  |  |
| $\theta_{2}$ | Hospitalisation with RSV affected by active infection with influenza | 1 | ${1/\psi}_{2}$ | Duration of RSV refractory period Rf (weeks) | 0,001 | 68 | <0 |  |  |
| ${1/\omega}_{2}$ | Duration of RSV residually positive period Rv (weeks) | 0,001 |  |  |  | 67 | <0 |  |  |
| ${1/\psi}_{2}$ | Duration of RSV refractory period Rf (weeks) | 0,001 |  |  |  | 67 | <0 |  |  |
| $\sigma_{1}$ | Transmission of influenza affected by active infection with RSV | 1 |  |  |  | 67 | <0 |  |  |
| ${1/\omega}_{1}$ | Duration of influenza residually positive period Rv (weeks) | 0,001 |  |  |  | 67 | <0 |  |  |
| ${1/\omega}_{1}$ | Duration of influenza residually positive period Rv (weeks) | 0,001 | ${1/\omega}_{2}$ | Duration of RSV residually positive period Rv (weeks) | 0,001 | 68 | <0 |  |  |
| $\sigma_{1}$ | Transmission of influenza affected by active infection with RSV | 1 | ${1/\omega}_{2}$ | Duration of RSV residually positive period Rv (weeks) | 0,001 | 68 | <0 |  |  |
| ${1/\omega}_{1}$ | Duration of influenza residually positive period Rv (weeks) | 0,001 | $\sigma_{1}$ | Transmission of influenza affected by active infection with RSV | 1 | 68 | <0 |  |  |
| $\sigma_{1}$ | Transmission of influenza affected by active infection with RSV | 1 | ${1/\psi}_{2}$ | Duration of RSV refractory period Rf (weeks) | 0,001 | 68 | <0 |  |  |

Supplementary Table 4. Start and end dates, size of each the catchment population for each age group a in the hospitalisation data, for each season of the study. Corresponding larger age groups b for the community ILI data are included in the bottom row.

| Season | Study start | Study end | Age groups (a) | | | |  |  |  | | |  | |  | |
| --- | --- | --- | --- | --- | --- | --- | --- | --- | --- | --- | --- | --- | --- | --- | --- |
|  |  |  | <6m | 6-12m | 1 | 2-4 | | 5-17 | | 18-49 | 50-64 | | 65-74 | | ≥75 |
| 10/11 | 2010-11-07 | 2011-03-19 | 0 | 0 | 0 | 0 | | 0 | | 569073 | 205408 | | 97797 | | 102896 |
| 11/12 | 2011-11-05 | 2012-04-13 | 9278 | 9278 | 19620 | 65457 | | 249490 | | 1047310 | 383442 | | 174625 | | 178095 |
| 12/13 | 2012-11-10 | 2013-04-20 | 5702 | 5701 | 12524 | 41393 | | 164894 | | 582931 | 226357 | | 103936 | | 107693 |
| 13/14 | 2013-11-08 | 2014-03-25 | 6219 | 6218 | 13633 | 45317 | | 191161 | | 664489 | 266113 | | 126169 | | 127139 |
| 14/15 | 2014-11-08 | 2015-03-26 | 9207 | 9207 | 19353 | 64785 | | 282133 | | 1088213 | 451687 | | 214489 | | 199975 |
| 15/16 | 2015-11-14 | 2016-04-30 | 4778 | 4778 | 9799 | 32638 | | 144939 | | 487534 | 209315 | | 100144 | | 94448 |
| 16/17 | 2016-11-08 | 2017-04-18 | 4574 | 4573 | 9656 | 30469 | | 145423 | | 473141 | 214647 | | 103094 | | 95766 |
| 17/18 | 2017-09-10 | 2018-06-26 | 4363 | 4363 | 9445 | 29435 | | 143414 | | 463322 | 215012 | | 104480 | | 95686 |
| 18/19 | 2018-09-08 | 2019-08-08 | 4327 | 4326 | 9366 | 30645 | | 149962 | | 479829 | 228595 | | 110380 | | 101302 |
| 19/20 | 2019-10-31 | 2020-03-12 | 4308 | 4307 | 8786 | 30024 | | 151048 | | 478337 | 234080 | | 110940 | | 103781 |
| 20/21 | 2020-12-10 | 2021-04-29 | 2865 | 2865 | 6239 | 20942 | | 111099 | | 346573 | 178397 | | 84113 | | 80844 |
| Community ILI age groups (b) | | | <5 | | | | | 5-14 | | 15-64 | | | ≥75 | | |

Supplementary Table 5. Clinical conditions used to identify admissions possibly associated with an influenza or RSV infection in the VAHNSI study, reproduced from ^25^.

| **Patients ≥5 years of age** | **ICD 9 Codes** | **ICD 10 Codes** |
| --- | --- | --- |
| Acute respiratory infection | 382·9; 460–466 | J00–J06, J20–J22, H66·90 |
| Acute myocardial infarction or acute coronary syndrome | 410–411 and 413–414 | I20–I25·9 |
| Asthma | 493–493·92 | J45·2–J45·22, J45·9–J45·998, J44–J44·9 |
| Heart failure | 428–429·0 | I50–I50·9; I51·4 |
| Pneumonia and influenza | 480–488 | J09–J18 |
| Chronic pulmonary obstructive disease | 490, 491, 492, 496 | J40–J44·9 |
| Myalgia | 729·1 | M79·1 |
| Metabolic failure (diabetic coma, renal dysfunction, acid-base disturbances, alterations to the water balance) | 250·1– 250·3; 584–586; 276–277 | E11·9, E10·9, E11·65, E10·65, E10·11, E11·01, E10·641, E11·641, E10·69, E11·00, E10·10, E11·69, N17·0, N17·1, N17·2, N17·8, N17·9, N18·1, N18·2, N18·3, N18·4, N18·5, N18·6M N18·9, N19, E87·0, E87·1, E87·2, E87·3, E87·4, E87·5, E87·6, E87·70, E87·71, E87·79, E86·0, E86·1 |
| Altered consciousness, convulsions, febrile-convulsions | 780·01–780·02; 780·09; 780·31–780·32 | R40·20, R40·4, R40·0, R40·1, R56·00, R56·01 |
| Dyspnea/respiratory abnormality | 786·0 | R06·0, R06–R06·9 |
| Respiratory abnormality | 786·00 | R06·9 |
| Shortness of breath | 786·05 | R06·02 |
| Respiratory abnormality NEC | 786·09 | R06·3, R06·00, R06·09, R06·83 |
| Respiratory symptoms/chest symptoms | 786·9 | R06·89 |
| Fever or fever unknown origin or non-specified | 780·6–780·60 | R50, R50·9 |
| Cough | 786·2 | R05 |
| Sepsis, systemic inflammatory response syndrome | 995·90–995·94 | R65·10, R65·11, R65·20, A41·9 |
| **Patients 0–4 years of age** | **ICD 9 Codes** | **ICD 10 Codes** |
| Acute upper or lower respiratory disease | 382·9; 460 to 466 | J00–J06, J20–J22 |
| Dyspnea, breathing anomaly, shortness of breath, tachypnea | 786·0; 786·00; 786·05–786·07; 786·09; 786·9 | R06·0, R06, R06·9, R06·3, R06·00, R06·09, R06·83, R06·02, R06·82, R06·2, R06·89 |
| Asthma | 493–493·92 | J45·2–J45·22, J45·9–J45·998, J44–J44·9 |
| Pneumonia and influenza | 480 to 488 | J09–J18 |
| Heart failure | 428–429·0 | I50–I50·9; I51·4 |
| Myalgia | 729·1 | M79·1 |
| Altered consciousness, convulsions, febrile convulsions | 780·01–780·02; 780·09; 780·31–780·32 | R40·20, R40·4, R40·0, R40·1, R56·00, R56·01 |
| Fever or fever unknown origin or non-specified | 780·6–780·60 | R50, R50·9 |
| Cough | 786·2 | R05 |
| Gastrointestinal manifestations | 009·0; 009·3 | A09·0; A09·9 |
| Sepsis, systemic inflammatory response syndrome | 995·90–995·94 | R65·10, R65·11, R65·20, A41·9 |
